# Virtual brain twins guide personalized treatment decision in schizophrenia

**DOI:** 10.64898/2026.05.06.26352533

**Authors:** Giacomo Preti, Huifang E Wang, Abolfazl Ziaeemehr, Marmaduke Woodman, Paula Prodan, Paul Triebkorn, Xiao Chang, Madalina Buciuman, Maria Sacha, Marius Fey, Martin Breyton, Viktor Sip, Gabriele Casagrande, Romain Guilhaumou, Pronia consortium, Amirhossein Esmaeili, Spase Petkoski, Long-Biao Cui, Jianfeng Feng, Egidio Ugo D’Angelo, Pierpaolo Sorrentino, Meysam Hashemi, Lia Domide, Damien Depannemaecker, Nikolaos Koutsouleris, Viktor Jirsa

## Abstract

Schizophrenia is a complex psychiatric disorder whose pathophysiology spans multiple spatial and temporal scales. Structural and functional neuroimaging studies have identified a broad range of disease-associated markers encompassing cortical atrophy, white matter disruptions, and aberrant functional connectivity patterns. Their application to personalized diagnosis and treatment selection has remained elusive. Here, we introduce the first Virtual Brain Twin (VBT) pipeline that integrates individual connectome-based network models with multimodal neuroimaging data, incorporating patient-specific structural connectivity, cortical thickness, and resting-state fMRI features to construct personalised whole-brain dynamical models. Dopaminergic and serotonergic signaling pathways are embedded within a mean-field framework, and simulation-based inference (SBI) is used to recover key pathophysiological parameters from individual patient data. The validity of this inference is first established using synthetic patients with known ground truth parameters, confirming that the pipeline can accurately identify underlying neurochemical states from simulated functional data. Applied to a cohort of 53 subjects in three clinical centers, the framework identifies personalized pathophysiological parameter regimes consistent with current neurobiological hypotheses of schizophrenia. Simulated pharmacological interventions within the VBT generate individualized medication effect trajectories that align retrospectively with known treatment outcomes, demonstrating the framework’s capacity to capture patient-specific pharmacological responses. These results establish a principled and extensible computational foundation for neuroimaging-guided personalized medicine in psychiatry, with direct implications for two prospective clinical trials conducted in Marseille and Munich as part of the Virtual Brain Twin project, designed to evaluate VBT-guided individualised antipsychotic treatment selection.

## 1 Introduction

Schizophrenia is a severe psychiatric disorder characterized by a spectrum of positive, negative, and cognitive symptoms (*1*), whose neurobiological substrates span multiple spatial and temporal scales (*2*). Extensive structural and functional neuroimaging studies have identified a broad range of disease-associated markers in affected individuals, encompassing cortical atrophy (*3*), white matter disruptions (*4*), and aberrant functional connectivity patterns (*5*). Despite progress in characterizing such markers at the group level, their translation into clinically actionable tools for guiding the diagnosis or treatment of individual patients remains critically limited. A central challenge lies in bridging the gap between population-level neuroimaging findings and the pronounced inter-individual variability that mirrors the clinical heterogeneity of schizophrenia. Addressing this challenge requires translational computational frameworks capable of integrating predictive models with multimodal neuroimaging data at the level of the individual patient. Here, we argue that Virtual Brain Twins (VBT), defined as personalized whole-brain models constructed from individual neuroimaging data, offer a principled framework for this purpose (*6, 7*).

Over the past two decades, machine learning approaches applied to structural and functional neuroimaging have demonstrated that generalizable signatures of schizophrenia exist at the single-subject level. Multivariate pattern analysis of structural MRI (sMRI) and resting-state functional MRI (fMRI) data have achieved a minimum accuracy of 60% (most of them 75–90%) for the diagnosis of schizophrenia (*8, 9*). Neuroimaging-based prediction has further revealed recoverable neurobiological subtypes within the heterogeneous construct of schizophrenia, including two distinct and reproducible neuroanatomical subtypes characterized respectively by widespread grey matter volume reduction and largely normal brain volumes (*10*). This challenges the notion that cortical volume loss is a universal feature of the disorder and suggests differential underlying aetiologies. Single-subject neuroimaging signatures have also shown promise in clinical high-risk states, suggesting that predictive biomarkers may be detectable prior to illness onset (*11*). These are significant achievements that establish the feasibility of single-subject neuroimaging-based prediction in psychiatry. However, these approaches share the fundamental limitation that they are only predictive but not mechanistic. They can identify who is likely to respond to treatment, but can neither explain why, nor can they simulate what would happen if a specific pharmacological intervention were applied to a patient at a specific dose. Critically, they do not provide an interpretable mapping between neuroimaging features and the underlying neurochemical parameters that drive both pathophysiology and treatment response. Rather than replacing these data-driven approaches, the Virtual Brain Twin framework presented here is designed to address this mechanistic gap by providing a biologically interpretable, causally grounded simulation of patient-specific brain dynamics and pharmacological response, opening a principled pathway from routine neuroimaging data to individualized treatment planning.

Schizophrenia is associated with well-characterized pathophysiological mechanisms, including dysregulation of dopaminergic and serotonergic signaling, that span multiple spatial scales, from receptor-level alterations to large-scale network dysfunction. Antipsychotic drugs exert their therapeutic effects by targeting these same receptor systems, with mechanisms that are increasingly well understood at the molecular and cellular level. We hypothesize that both the pathophysiological alterations and the pharmacological effects of antipsychotic medications translate consistently to the level of whole-brain network dynamics, such that they can be captured and predicted within a personalized connectome-based modeling framework. If this holds, then individual connectome-based virtual brain modeling (*12*) can provide a principled and tractable framework for predicting patient-specific changes in brain activity as a function of drug type and dosage. To test this hypothesis, we first develop the theoretical foundations of connectome-based network models, incorporating dopaminergic, serotonergic, excitatory, and inhibitory signaling pathways within a mean-field dynamical framework. We then apply this framework to cohorts comprising 53 schizophrenic patients and healthy controls across three clinical centres, validating VBT predictions against empirical neuroimaging data and longitudinal clinical outcomes. Local neural dynamics are modeled using a mean-field approximation, with inter-regional interactions mediated through excitatory, inhibitory, dopaminergic, and serotonergic pathways derived from patient-specific diffusion MRI tractography and literature-derived receptor density maps. Personalization further incorporates cortical thickness estimates from T1-weighted MRI and optimizes 20 out of 72 resting-state fMRI data features summarizing both activation and dynamic functional connectivity. A simulation-based inference (SBI) algorithm (*13, 14*) then recovers three key pathophysiological parameters from individual patient data, namely cortical dopaminergic drive, subcortical dopaminergic drive, and whole-brain serotonergic drive, where drive denotes the net modulatory influence of a neuromodulatory system on its target circuits. The pharmacological action is modeled by modulating a drug-specific variable to simulate receptor-level interactions in D_1_, D_2_, and 5-HT_2*A*_, allowing for the in silico prediction of dose-dependent effects of pharmacological interventions, yielding patient-specific modulation profiles for each medication.

This work introduces the first personalised VBT pipeline for schizophrenia, integrating subject-specific structural connectivity, cortical thickness, and resting-state functional features within a unified inference and medication simulation framework. It establishes synthetic patients as a principled validation tool for parameter recovery, providing ground truth benchmarks that are inaccessible in real clinical data. It demonstrates, for the first time, that in silico pharmacological simulation within a personalised whole-brain model can retrospectively capture differential treatment responses at the individual patient level. Together, it establishes an avenue for VBT-guided personalized medicine in psychiatry.

## 2 Results

### 2.1 Translating pathophysiological and pharmacological mechanisms into virtual brain twins

Schizophrenia is characterized by an imbalance between the mesolimbic and mesocortical dopamin-ergic pathways. Two complementary mechanisms are now well established: subcortical hyperdopaminergia, wherein excessive dopamine activity at D_2_ receptors in the striatum drives positive symptoms; and cortical hypodopaminergia, where insufficient dopamine signaling at D_1_ receptors in the prefrontal cortex underlies negative and cognitive symptoms (*15*). Serotonin serves as a critical modulator of dopaminergic activity. Specifically, 5-HT_2*A*_ receptors, localized on glutamatergic neurons and dopaminergic terminals, are often hyperactive in schizophrenia, further exacerbating the dopaminergic dysregulation (*16*). The VBT framework incorporates these neuro-chemical dynamics by explicitly modeling the synergistic effects of dopamine and serotonin (Fig. 1A). We distinguish between subcortical and cortical dopaminergic effects by introducing two independent scaling parameters 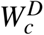, 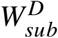, along with *W*^*S*^ for serotonergic effects, allowing for the representation of the distinct pathological states across brain regions. Throughout this work, 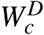 denotes the cortical dopaminergic drive (defined as the net modulatory influence of dopaminergic projections on cortical circuits), 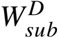 denotes the subcortical dopaminergic drive, and *W*^*S*^ the global serotonergic drive, representing a whole-brain serotonergic modulation parameter. Accordingly, the structural connectivity was classified into four neurotransmitter-specific networks: glutamatergic, GABAergic, dopaminergic, and serotonergic, by integrating patient-specific white matter tractography derived from diffusion MRI with established neuroanatomical knowledge of projection directionality and neurotransmitter density (Fig. 1B).

**Figure 1:**
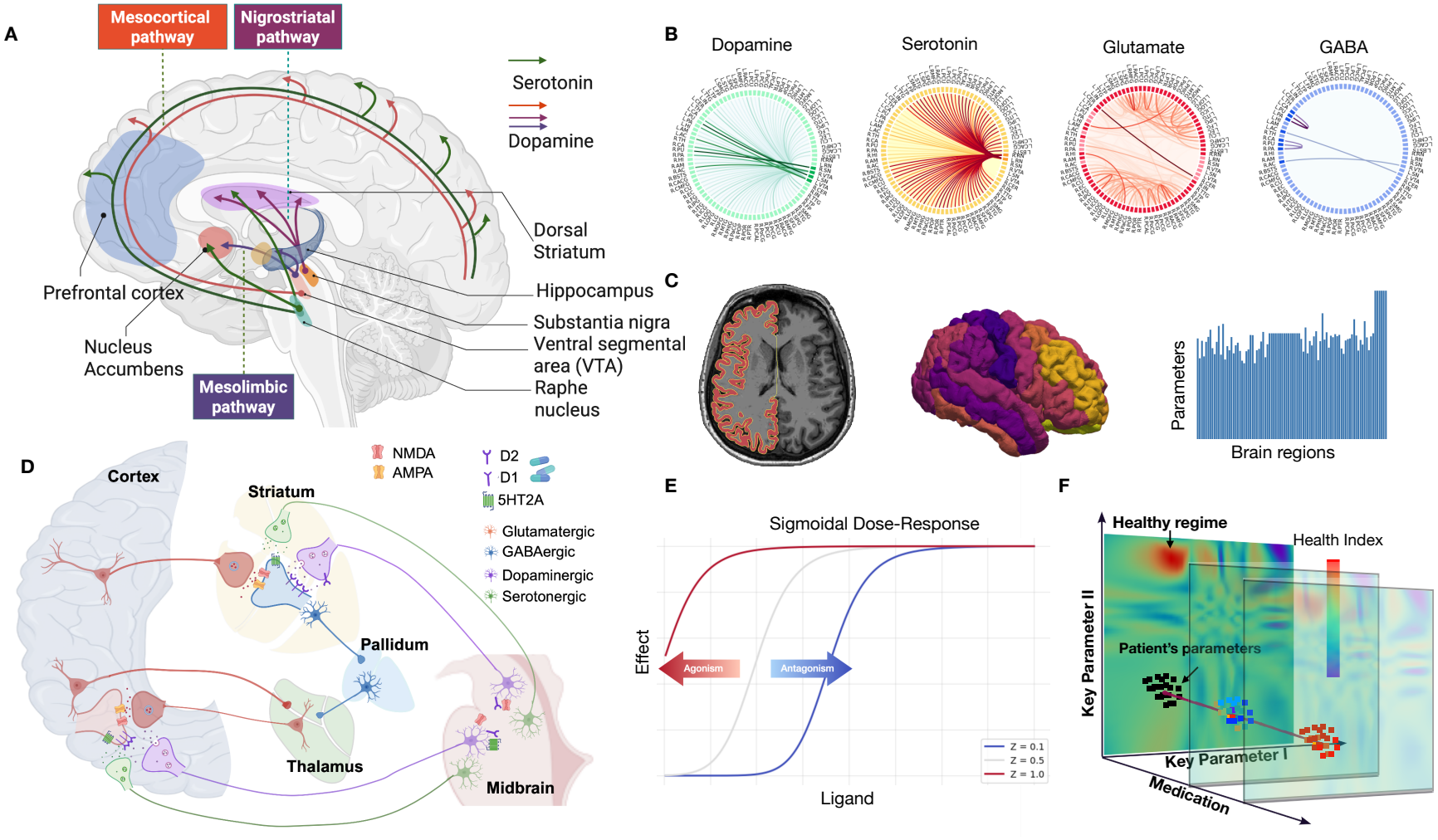
Translating pathophysiological and pharmacological mechanisms of schizophrenia into Virtual Brain Twins. (**A**) Three major neuromodulatory pathways: mesolimbic, mesocortical, and nigrostriatal, for dopamine (DA) and serotonin (5-HT), highlighting key brain regions including the VTA, substantia nigra, raphe nuclei, striatum, nucleus accumbens, and prefrontal cortex. (**B**) Structural connectivity classified into four neurotransmitter systems: dopaminergic, serotonergic, glutamatergic, and GABAergic, illustrated for a representative patient. (**C**) Cortical thickness visualized across the cortical surface (left) and quantified per brain region (middle), subsequently mapped to a key parameter within the Virtual Brain Twins framework (right). (**D**) Major antipsychotic drugs primarily target dopamine D_1_ and D_2_ receptors and the serotonin 5-HT_2*A*_ receptor, which are abundantly expressed in the cortex, striatum, and midbrain structures, including the substantia nigra, ventral tegmental area, and raphe nuclei. To illustrate the underlying synaptic mechanisms, we provide magnified views of representative synaptic configurations in two functionally distinct regions: the striatum and the prefrontal cortex, highlighting the principal neurotransmitters and their associated receptor subtypes at each site. (**E**) Sigmoidal dose-response curve as a function of ligand concentration modeling pharmacological effects of antipsychotic medications; receptor antagonism produces a rightward shift, indicating that higher ligand concentrations are required to achieve equivalent receptor occupancy. (**F**) Within the Virtual Brain Twins parameter space, a healthy regime is defined from biophysiological knowledge derived from healthy subjects with healthy structural connectivity and cortical thickness, i.e., characterized by higher cortical and lower subcortical dopamine. Individual patient parameter distributions (black dots) are inferred from patient-specific both structural and functional data, and a health index (color scale) is derived from functional MRI features. Along the medication axis, functional MRI features are computed across doses according to each drug’s pharmacological mechanism, with therapeutic efficacy determined when the health index surpasses a predefined threshold.

Beyond neurochemical dysregulation, schizophrenia is characterized by significant structural alterations, most notably accelerated cortical thinning and reduced grey matter volume, as observed in neuroimaging studies (*17–20*). These structural alterations are increasingly recognized as proxies of specific disease’s pathophysiologyical processess (*17*). For example, volume loss might index synaptic changes, altered myelination, changes in neuronal soma size, and dendritic modifications (*21*). Decreased synaptic markers have been reported in the hippocampus, the anterior cingulate cortex, and the prefrontal cortex (*22*), which are plausibly linked to the cortical volume reduction and thinning (*17, 21*). Moreover, if cortical volume loss and cortical thinning is due to either a reduced number of neurons or neuropil space, this is likely to be accompanied by lower synaptic activity. Since the majority of cortical neurons are glutamatergic pyramidal cells, it can be arguably assumed that cortical volume loss can reflect, at least in part, a loss in glutamatergic activity. It has been repeatedly shown that the dendrites, axons, cell bodies, and levels of synaptophysin, a marker of axon boutons of cortical glutamatergic neurons, are reduced in size in schizophrenia (*23*). As a consequence of the reasoning above, the parameter *J*_*a*_ (Eq. 5) scales self-excitation within a given region and *J*_*sa*_ its excitatory input from other brain regions. These parameters were identified as being linked to cortical thickness (Fig. 1C), motivating the introduction of subject-specific regional variability into the model. Specifically, these two parameters vary across brain regions according to individual cortical gray matter thickness, as described below. Regional cortical gray matter thickness was estimated for each brain region using the FreeSurfer preprocessing pipeline applied to subject-specific T1-weighted MRI data. To account for normative developmental and morphological variability, these regional measurements were compared against a population-based reference model derived from the CENTILE framework (*24*), which provides lifespan normative estimates of regional cortical thickness stratified by age and brain size. The regional parameters *J*_*a*_ and *J*_*sa*_ were then mapped as a function of the deviation of each subject’s cortical thickness from the normative mean, expressed in units of the normative standard deviation, after controlling for age, sex, and total brain size.

Antipsychotic drugs exert their therapeutic effects primarily through agonism or antagonism of dopamine D_1_ and D_2_ receptors and the serotonin 5-HT_2*A*_ receptor, which are differentially expressed across cortical, striatal, and midbrain circuits (Fig. 1D). At the synaptic level, these receptor systems operate through distinct mechanisms depending on the local circuit architecture. Dopaminergic signaling in the striatum is predominantly mediated through D_1_ and D_2_ receptor modulation of GABAergic medium spiny neurons, while serotonergic inputs via 5-HT_2*A*_ receptors further modulate striatal activity by regulating dopamine release and GABAergic interneuron excitability, whereas in the prefrontal cortex, the interplay between dopaminergic and serotonergic inputs critically regulates glutamatergic pyramidal cell activity (Fig. 1D). Most antipsychotic drugs act as reversible, competitive antagonists at dopaminergic D_1_ and D_2_ receptors, or serotonergic 5-HT_2*A*_ receptors. Here, “reversible” indicates that the drug does not permanently bind to the receptor. “Competitive” means that the drug competes with the endogenous neurotransmitter (dopamine or serotonin) for the same binding site, thereby reducing the receptor’s effective affinity for its ligand. As a consequence, higher concentrations of neurotransmitter are required to achieve the same receptor activation when antagonist levels increase (*25*). In the VBT, the influence of a reversible, competitive antagonist is captured by shifting left or right a sigmoidal dose-response curve that models the effect of the activation of the receptors on the electrical activity of each brain region, as shown in Fig. 1E.

Medication effects are introduced by modulating the dose-response curve, which encodes the pharmacological action of antipsychotic drugs on D_1_, D_2_, and 5-HT_2*A*_ receptors, with the therapeutic goal of displacing the patient’s brain state toward a healthy regime defined by functional MRI (fMRI) features (Fig. 1F). This healthy regime is established from both neurophysiological knowledge and empirical data derived from healthy subjects with normative structural connectivity and cortical thickness. As an illustrative example in a two-dimensional parameter space, cortical dopamine drive is characteristically higher in the healthy regime relative to the schizophrenic regime, whereas subcortical dopamine levels can be elevated in schizophrenia as compared to healthy subjects. Within this parameter space, a health index is defined from fMRI-derived functional features, providing a continuous scalar measure of proximity to the healthy regime (Fig. 1F, color scale). Patient-specific key parameters are obtained by fitting VBT models to individual structural and functional data through Bayesian inference. Medication is then simulated by variations along the pharmacological axis, where each drug’s mechanism of action determines the trajectory through parameter space. Functional MRI features are computed across a range of doses, and therapeutic efficacy is determined when the health index surpasses a predefined threshold, providing a principled, patient-specific framework for in silico drug prediction and optimization.

### 2.2 The workflow of virtual brain twins in schizophrenia

The VBT for schizophrenia (shown in Fig. 2) are constructed upon a whole-brain network comprising 86 regions defined by the Desikan–Killiany (DK) atlas (*26*). Brain region parcellation is derived from subject-specific T1-weighted MRI data, while structural connectivity is obtained from diffusion-weighted MRI using tractography. Connectivity masks are used to distinguish white matter fiber tracts associated with excitatory, inhibitory, dopaminergic, and serotonergic pathways. Neural dynamics at each brain region are modeled using mean-field modeling (Eq. 1-6) incorporating neuromodulatory effects, explicitly accounting for dopamine and serotonin influences (Eq. 7-11) (Fig. 2A). Regional cortical thickness extracted from T1-weighted MRI is mapped to the regional parameters *J*_*a*_ and *J*_*sa*_, under the hypothesis that regions with larger cortical thickness exhibit stronger self-excitation exert stronger excitatory influences on other regions. Neuromodu-latory receptor density distributions are incorporated from cohort-level PET data, considering D_1_, D_2_, and 5-HT_2*A*_ receptors (Fig. 2B).

**Figure 2:**
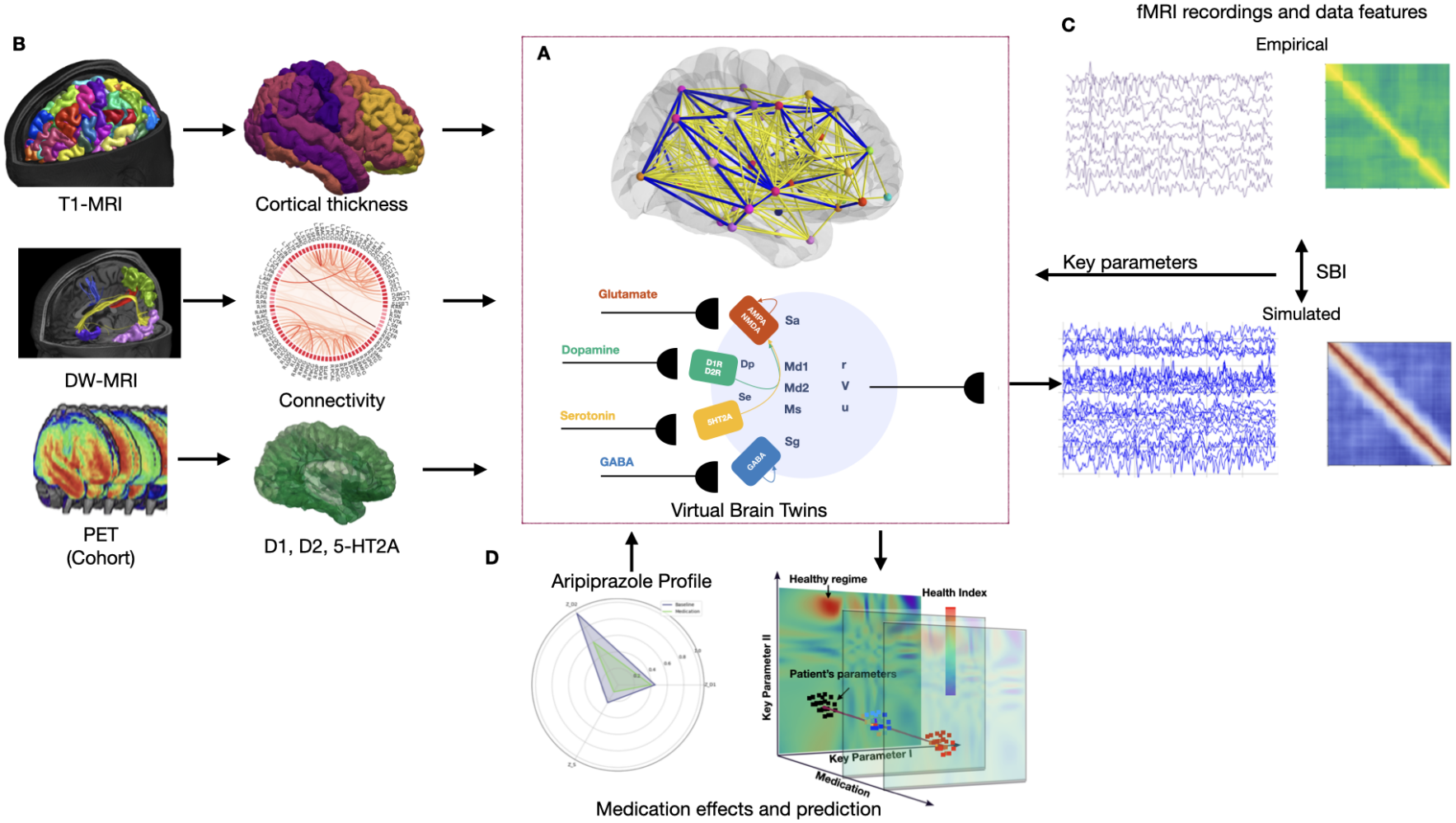
The Virtual Brain Twins clinical trial pipeline for schizophrenia. (**A**) Virtual Brain Twin is constructed as a personalized whole-brain network model derived from patient-specific multimodal neuroimaging data. The network comprises nodes representing individual brain regions and edges encoding their structural interconnections. At each node, a mean-field model captures local neural dynamics, incorporating the contributions of glutamatergic, dopaminergic, serotonergic, and GABAergic signaling, as well as the network-mediated input from all connected regions. (**B**) Regional node properties are derived from T1-weighted MRI, which defines the anatomical atlas and provides cortical thickness estimates used to introduce regional heterogeneity via the parameter *J*_*a*_. Structural connectivity is obtained from the topological organization of white matter pathways extracted through diffusion-weighted MRI tractography. Neuromodulatory receptor distributions, including dopamine D_1_, D_2_, and serotonin 5-HT_2*A*_ receptors, are informed by population-level PET receptor density data. (**C**) The VBT generates synthetic fMRI data and associated functional data features, which are used to train a deep neural density estimator within the simulation-based inference (SBI) framework. By fitting the trained SBI model to patient-specific empirical fMRI recordings, this Bayesian approach recovers the posterior distributions of three key neuromodulatory parameters: cortical dopaminergic drive, subcortical dopaminergic drive, and global serotonergic drive. (**D**) Pharmacological interventions are introduced into the model through drug-specific neuromodulatory profiles (illustrated here for aripiprazole) enabling in silico prediction of individual treatment response and therapeutic efficacy.

The resulting whole-brain models are used to simulate fMRI data and compute relevant data features. A simulation-based inference (SBI) framework is trained on features extracted from simulated data and subsequently applied to estimate three key neuromodulatory parameters, 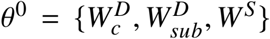, from empirical fMRI recordings (shown in Fig. 2C). This process yields individualized virtual brain twins, whereby personalized model parameters are inferred from patient-specific functional neuroimaging data.

Pharmacological interventions are introduced into the model through drug-specific neuromodulatory profiles. Each drug differently modulates the parameter Z that controls the shift of the sigmoidal functions (Eq. 9-11), according to its mechanism of action (competitive antagonist or partial agonist) and its receptor binding affinity (Ki). Increasing doses of the drug increase the occupancy of the receptor, that translates into a shift of the sigmoidal functions. For each receptor, the corresponding Z is increased or decreased by scaling coefficients that are determined by the Ki of the modeled drug for that particular receptor. A pharmacokinetic–pharmacodynamic framework described by (*27*) enables the conversion of the receptor occupancy (and therefore the simulated shift of Z) in clinically interpretable units of mg/day. In Fig. 2D, we show an example of how the three (Z) parameters are rescaled following the administration of an illustrative dose of aripiprazole. In the radar plot, the original values of the Z parameters in the absence of medication are represented by the purple triangle, while the green triangle represents the resulting values after rescaling to account for the selected dose. Predicted fMRI data are then generated using virtual brain twins by integrating the personalized parameter *θ*^0^ with the medication parameter *Z*. Therapeutic efficacy is quantified through a medication efficacy score *E*_*D*_, defined by comparing simulated fMRI features (listed in Table S3) in the medicated state against those of the baseline unmedicated state. The SBI approach allows us to calculate the posterior probability *P*(*E*_*D*_ | *θ*^0^, *Z*), providing a patient-specific prediction of individual treatment response.

### 2.3 Synthetic Patients

We use synthetic patients to afford full experimental control over key neurochemical parameters and to establish a known ground truth, neither of which is achievable in vivo. Synthetic patients were constructed from the anatomical data of 9 subjects, from which cortical thickness, brain par-cellations, and structural connectivity were derived. Following a systematic parameter exploration of noise and excitability weights *W*^*e*^, these two parameters were fixed as optimal working points for all subsequent analyses (Supplementary Fig. 2). We then used SBI to infer the joint posterior distribution of the key parameters defined by 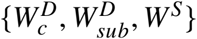 corresponding to cortical dopaminergic, subcortical dopaminergic and serotonergic drives, respectively. This Bayesian approach allowed us to precisely define the neurochemical state of the synthetic patients and directly link these states to emergent fMRI features.

The 72 data features (Supplementary Table 1) were derived from fMRI-BOLD signals across each set of model parameters, with the Amplitude of Low Frequency Fluctuations (ALFF), Functional Connectivity (FC), and Functional Connectivity Dynamics (FCD) shown as examples in Fig. 3A. Three representative features, Global Brain Coupling (GBC), defined as the mean whole-brain Functional Connectivity matrix, Fluidity (the variance of the FCD), and the mean FC between the Left Putamen and all other brain regions, are illustrated in the 3D parameter space spanned by 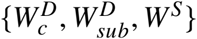 (Fig. 3B), presented as both full three-dimensional representations and one-dimensional marginal projections. The distributions of these features across all parameter combinations are shown alongside their dependence on each parameter individually (Fig. 3B). The serotonergic drive *W* ^*S*^ and cortical dopaminergic drive 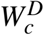 both exert a marked influence on GBC and Fluidity. By contrast, GBC and Fluidity shows minimal sensitivity to the subcortical dopaminergic drive 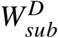, which instead modulates the mean FC of the Left Putamen in a step-like increasing way.

**Table 1.** Summary of posterior distribution comparisons, Kolmogorov–Smirnov (KS) distances, two-sided independent-samples t-tests, and SBI shrinkage for the three inferred parameters in the FES and HC groups. The mean difference is defined as FES − HC.

| **Parameter** | **Mean difference [95% CI]** | *P*(FES > HC) | **KS distance** | **t-test p-value** |
|---|---|---|---|---|
| *W*<sup>*S*</sup> | −0.148 [−0.545, 0.268] | 0.247 | 0.130 | 0.638 |
| *W*<sub>*c*</sub><sup>*D*</sup> | −0.864 [−1.386, −0.330](†) | 0.001 (#) | 0.448 | 0.260 |
| *W*<sub>str</sub><sup>*D*</sup> | 0.158 [−0.318, 0.520] | 0.768 | 0.315 | 0.653 |

**Figure 3:**
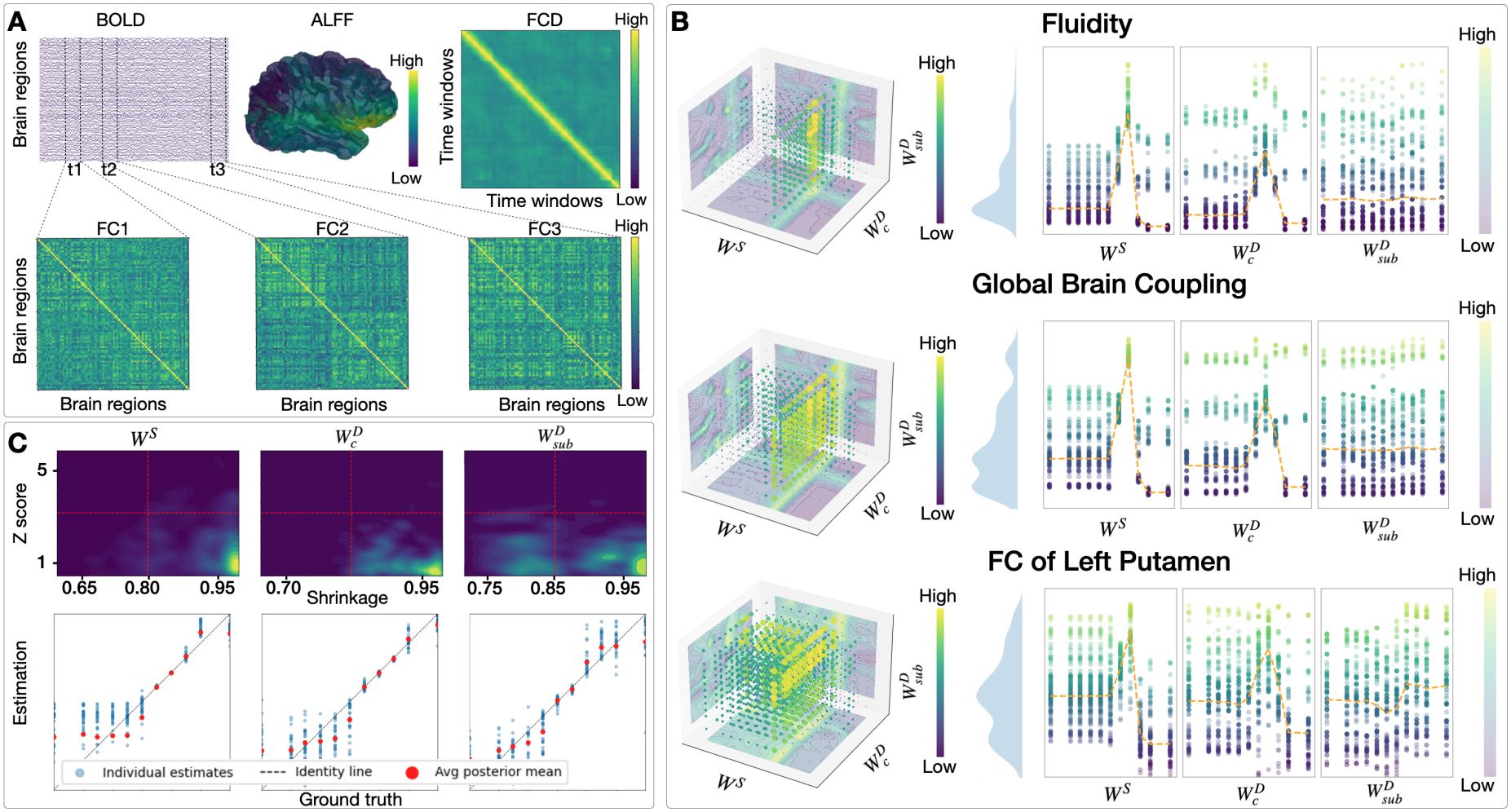
Data features of synthetic patients in parameter space and validation of key parameter inference. (**A**) Simulated BOLD signals generated by a specific set of model parameters and the resulting data features. Top row: fMRI time series from randomly selected regions, the amplitude of low-frequency fluctuations (ALFF) mapped onto the 3D brain, and functional connectivity dynamics (FCD). Bottom row: Functional connectivity matrices across three selected time windows. (**B**) Mapping of data features across three model parameters: serotonergic coupling, cortical dopaminergic coupling, and striatal dopaminergic coupling. Three data features are examined: global brain coupling (GBC), Fluidity, and mean FC of the Left Putamen. For each feature (rows): (Left) 3D scatter plots illustrate the explored parameter space, where each point represents a unique parameter combination and is color-coded by the corresponding feature value. (Middle) Violin plots show the distribution of feature values across the full parameter sweep. (Right) Scatter plots depict the relationship between each feature (y-axis) and each individual parameter (x-axis), marginalizing over the remaining parameters. The dashed orange line indicates the feature mean. (**C**) Validation of simulation-based inference (SBI) results. The top row displays kernel density estimate (KDE) heatmaps for each inferred parameter. The y-axis represents the posterior z-score and the x-axis shows the shrinkage. Lower posterior z-scores and higher shrinkage values (closer to 1) indicate a well-calibrated posterior concentrated around the true parameter values. The bottom row shows scatter plots comparing true parameter values (x-axis) with inferred estimates (y-axis). Points closer to the identity diagonal indicate higher accuracy. Red markers denote the mean of the posterior distribution.

We used SBI to infer the posterior distributions of key parameters from functional data features extracted from simulated BOLD signals. For synthetic patients, the parameters used for simulation serve as ground truth. Within the SBI, a deep neural density estimator was trained on 11^3^ parameter combinations, each yielding a probabilistic mapping between the parameters and set of extracted functional features. The parameter ranges were selected based on the parameter-versus-feature plots (Fig. 3B), which showed that the model features respond to changes in the parameters within the selected ranges, while values outside these ranges produce negligible changes in the extracted features. We evaluated the calibration of the estimator training by splitting the 11^3^ parameter combinations and their corresponding data features into training and test sets. The estimator was trained on the training set, and its performance was subsequently evaluated on the held-out test set. Specifically, the trained estimator was provided with the data features associated with the test parameter combinations, and its ability to recover the corresponding parameters was assessed by computing the discrepancy between the ground-truth parameters and their posterior estimates, expressed in z-score units. We additionally quantified posterior shrinkage, a measure of reduction in posterior uncertainty relative to the full parameter range. Fig. 3C (top) shows that most parameter combinations yield high shrinkage and small posterior z-scores, indicating well-calibrated and accurate inference. Fig. 3C (bottom) further demonstrates that the posterior distributions of 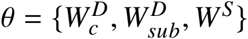 successfully recover the ground truth values, confirming that SBI can reliably identify the underlying neurochemical states from simulated functional data.

We next explored how different brain states emerge from systematic variations of either patho-physiological parameters 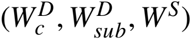 or pharmacological parameters (*Z*). By varying these parameters, we generate simulated neural activity from which synthetic data features are extracted, enabling in silico exploration of disease mechanisms and drug effects. To model schizophrenia-related alterations, we focused on parameter regimes consistent with current hypotheses, namely reduced cortical dopaminergic drive, increased cortical serotonergic, and elevated subcortical dopaminergic drive. Fig. 4A shows three representative conditions. The healthy state (light green) is characterized by balanced neurotransmitter activity. The pathological condition (purple) reflects decreased cortical dopaminergic, increased subcortical dopaminergic, and increased cortical serotonergic drive. The medicated condition (green) retains the same pathophysiological parameters as the pathological state, as pharmacological effects are modeled exclusively through modulation of the *Z* parameters rather than changes in the underlying disease pathophysiological parameters. The effect of medication is illustrated in Fig. 4B using radar plots. These show how three example drugs modulate the parameters 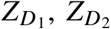, and *Z*_*S*_, which control the position of the sigmoidal transfer functions (Eq. 9–11). Starting from baseline values (purple triangles: before medication), the parameters are decreased or increased according to a dose-dependent factor 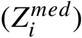, while preserving their relative ratios determined by receptor affinities (see Table 8 in the Methods section). By progressively varying 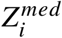 across increasing simulated dose levels *i*, we model the dose-dependent effects of the drug on the three neuromodulatory systems and, consequently, on brain activity. Changes in parameter space translate into systematic shifts in data feature space, illustrated by the two example feature spaces in Fig. 4C. The first shows GBC versus Fluidity, capturing global network properties. The second shows the FC between the Left Caudate nucleus and the Left Hippocampus, versus the FC between the Left Putamen and the left Caudal Anterior Cingulate cortex, reflecting region-specific connectivity. Small purple dots represent simulations obtained by exploration of the pathophysiological parameters 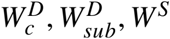. Within this space, the large light-green point corresponds to the healthy condition, while the large purple point represents the pathological state. The continuous trajectory between them illustrates how progressive alterations in neurotransmitter balance drive the system from a healthy to a pathological state.

**Figure 4:**
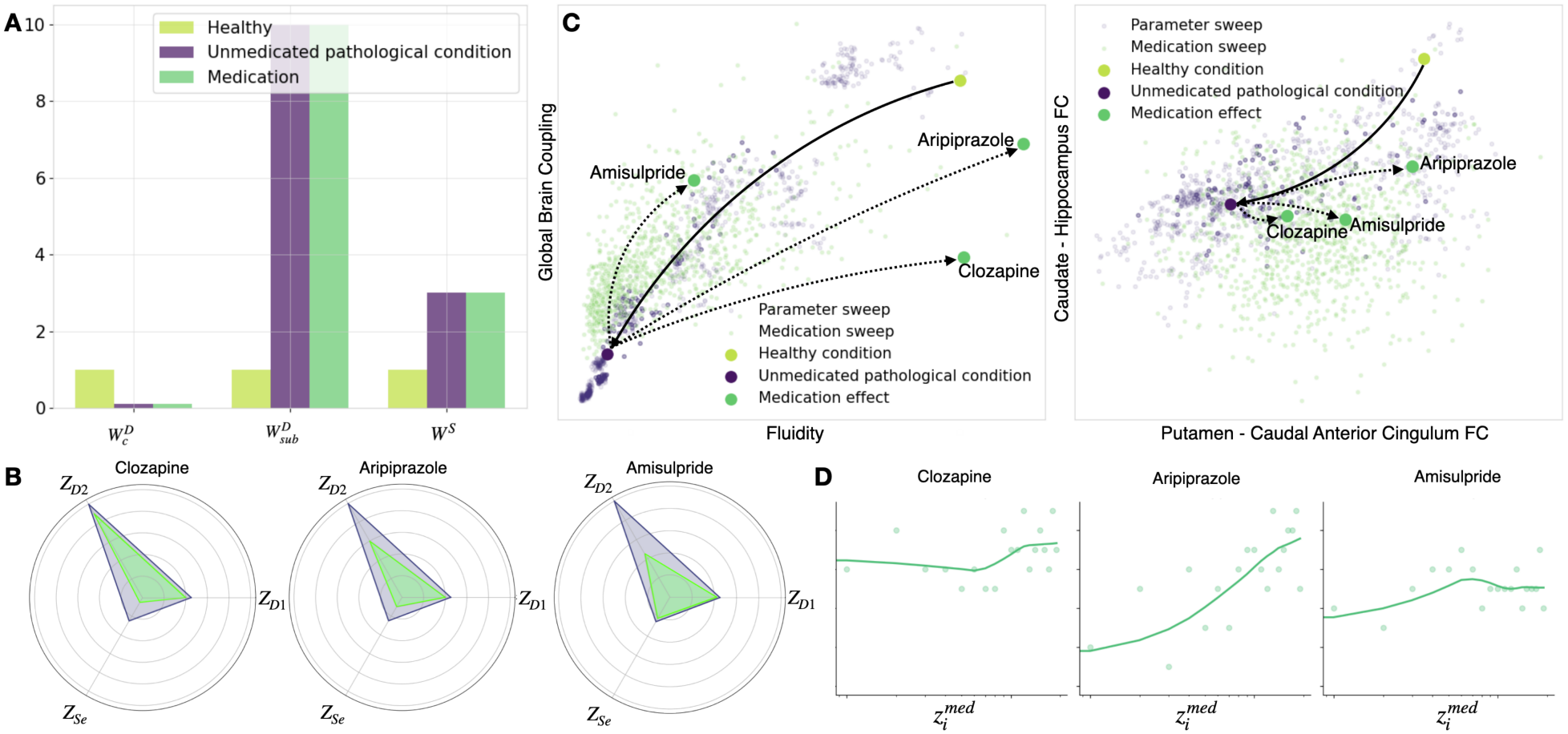
Medication effects simulated on the synthetic patients. (**A**) Pathophysiological parameter sets. Three examples of simulated states are driven by changing three key pathophysiological parameters. The light-green columns represent a healthy state, characterized by low serotonin drive, high cortical dopamine drive, and low subcortical dopaminergic drive. The purple columns represent a pathological state consistent with current hypotheses of schizophrenia: increased cortical 5-HT_2*A*_ activity, reduced cortical dopamine, and elevated subcortical dopaminergic coupling. The green columns on the right represent the same patho-physiological parameter sets as the disease condition — unchanged by medication — since these parameters describe the underlying pathophysiology rather than the pharmacological intervention itself. (**B**) Medication effect profiles on model parameters. Radar plots illustrate the effect of three example drug doses on the scaling parameters Z of the sigmoidal dose-effect functions (Eq. 9–11), compared to baseline values (purple triangles). Green triangles indicate how Z shifts under a given drug dose, illustrating the pharmacological modulation of each model parameter. (**C**) Feature space representation of disease and treatment effects. Two example data feature spaces are shown: Global Brain Coupling vs. Fluidity (left), and Putamen–Caudal Anterior Cingulate FC vs. Caudate–Hippocampus FC (right). Small purple and green dots represent simulation outputs across the full range of pathophysiological and pharmacological parameters, respectively. The large light-green dot corresponds to the healthy condition, while the large purple dot represents the pathological (unmedicated) condition defined in panel A. The displacement from the healthy to the pathological point illustrates how changes in pathophysiological parameters shift the patient’s position in data-feature space. From the pathological state, three dashed arrows show how simulated data features evolve under increasing doses of amisulpride, aripiprazole, and clozapine, capturing how pharmacological modulation moves the system through feature space toward (or away from) the healthy regime. (**D**) Dose–response relationships. Dose–response curves show medication effectiveness scores (y-axis) as a function of drug dose 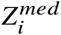 (x-axis), computed from simulated features. Clozapine achieves a high effectiveness score that plateaus across doses, while aripiprazole shows a clear dose-dependent increase. Amisulpride remains the least effective across all doses.

From the pathological condition, pharmacological perturbations are introduced by 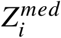. The resulting simulations are shown as small green points in data feature space 4C. For each drug, a representative dose is highlighted (large green points), corresponding to the parameter shifts illustrated in the radar plots Fig. 4B. The dashed trajectories indicate how each medication moves the system through data feature space, reflecting distinct degrees of recovery towards the healthy regime. Finally, Fig. 4D summarizes the dose–response relationships for the three medications. The medication effectiveness score *E*_*D*_, computed from the simulated functional data features, is plotted as a function of the scaling factor 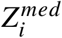 for increasing simulated doses *i*. Clozapine achieves a high effectiveness score that remains relatively stable across doses, suggesting that even low doses may produce substantial therapeutic benefit. Aripiprazole exhibits a clear dose-dependent increase in effectiveness, indicating that higher doses progressively improve outcomes. Amisulpride shows the weakest overall response across all doses examined. Together, these results demonstrate that the model captures differential drug efficacy and provides a quantitative framework for comparing pharmacological interventions in silico. Notably, comparable therapeutic outcomes may be achievable with either low doses of clozapine or higher doses of aripiprazole, offering potentially useful guidance for individualised treatment selection.

### 2.4 Example Patients

To validate our pipeline for medication prediction, we first applied the VBT framework to a cohort of 13 patients for whom resting-state fMRI data were available both before and after medication. Nine patients were drawn from the PRONIA dataset (Ludwig-Maximilians-University, Munich, Germany) and were treated with aripiprazole, while four patients were obtained from another dataset from Xijing Hospital (Xi’an, China) with four mixed pharmacological treatments. Here, we present results from three representative patients: one who responded to aripiprazole, one who did not respond, and one who responded to olanzapine. For each patient, we illustrate the key steps of the pipeline and its final medication response prediction.

#### 2.4.1 Patient 1: Aripiprazole responder

First, we reconstructed the virtual brain twin for each patient using individual anatomical data — T1-weighted MRI and diffusion-weighted MRI — to inform parameters related to cortical thickness and structural connectivity. Pre-medication resting-state fMRI recordings were preprocessed using fMRIPrep (*28*), and nuisance signals were removed using aCompCor regressors (*29*). From the cleaned BOLD time series, we computed regional ALFF, the whole-brain FC matrix, and FCD obtained via sliding-window Pearson correlations (shown in Fig. 5A) from which we retrieved a total of 72 features, including GBC, Fluidity, and selected regional FC and ALFF metrics (see Supplementary Table 1).

**Figure 5:**
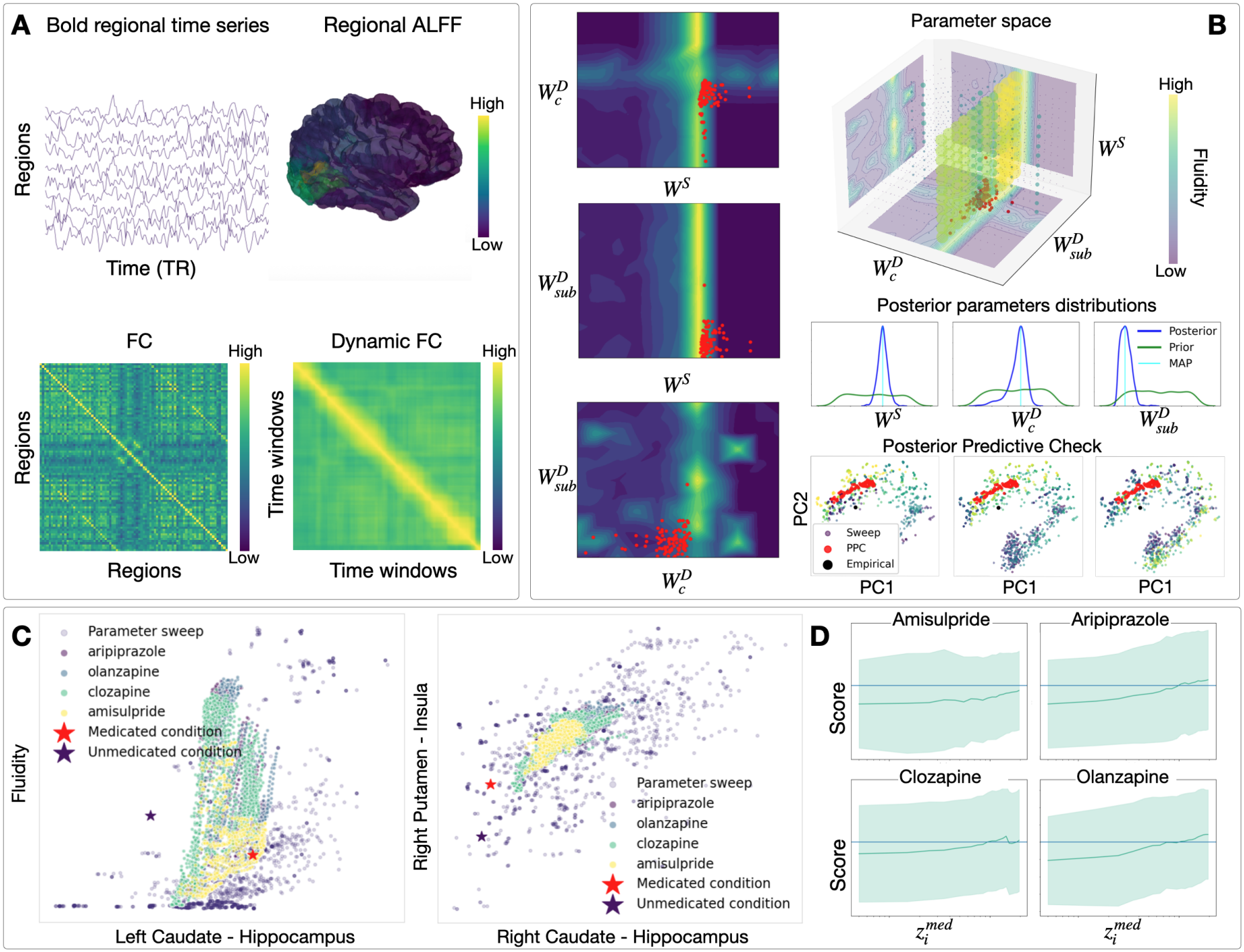
Patient 1: VBT analysis of a treatment responder to aripiprazole. (**A**) The patient’s resting-state fMRI BOLD signal and three data features: regional ALFF mapped onto the 3D brain volume, the FC matrix computed over the entire recording, and the FCD matrix. (**B**) Posterior distributions of key pathophysiological parameters inferred by SBI, shown in both parameter space and data feature space. (Right middle) Posterior distributions of the three key pathophysiological parameters — serotonergic drive *W*^*S*^, cortical dopaminergic drive 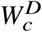, and subcortical dopaminergic drive 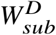 — identified by SBI as best matching the patient’s empirical data. The green curve represents the prior distribution (the full initial parameter space), while the blue curve shows the posterior inferred from the patient’s data. The light blue line marks the maximum a posteriori (MAP) estimate. The inferred posterior parameter estimates (red points) mapped onto the parameter space, shown as a 3D scatter plot (Middle top) and as 2D pairwise projections marginalising over the third parameter (Left). The background colour is scaled according to fluidity as a representative data feature. Data features extracted from simulations generated using the posterior parameter distributions (red points), also shown in the space of the first two principal components (PCA) (Middle bottom). Each subfigure is coloured by the corresponding parameter values from the parameter sweep. The black point represents the patient’s empirical data. (**C**) Medication effects in pairwise data feature space. Two example feature spaces are shown: Left Caudate–Hippocampus FC versus Fluidity, and Right Putamen-Insula FC versus Right Caudate–Hippocampus FC. Smaller dots represent simulations from both pathophysiological and pharmacological parameter sweeps. Pharmacological simulations include four medications — clozapine, aripiprazole, amisulpride, and olanzapine — at increasing doses, colour-coded by medication type. The patient’s empirical data are shown as a purple star (baseline, before medication) and a red star (follow-up, after administration of aripirpazole). (**D**) Medication effectiveness scores for each drug across increasing doses. Amisulpride shows a gradually increasing response that never reaches the threshold, indicating ineffectiveness at all tested doses. Aripiprazole and olanzapine exhibit a clear dose-response relationship that crosses the threshold, while clozapine reaches peak effectiveness at intermediate doses before its effect diminishes.

Using the subject-specific structural connectome, we performed a parameter exploration of 11^3^ simulations across three pathophysiological parameters (serotonergic drive *W*^*S*^, cortical dopaminergic drive 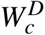, and striatal dopaminergic drive 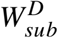). Each simulation lasted 5 minutes, and a hemodynamic forward model was applied to transform simulated neural activity into BOLD signals. From these simulations, we extracted the same 72 data features, resulting in a set of 11^3^ × 72 configurations. This dataset was used to train deep neural density estimators in the SBI approach, which approximates the posterior distribution directly from simulated parameter-feature pairs using normalizing flows. The trained model was then used to infer the posterior parameter distributions that best explain the empirical data (Fig. 5B). It is possible to visualize the posterior distribution of the three inferred parameters (red points) in both 3D parameter spaces and 3 pair-wise 2D parameter spaces, where the background is colored according to an example data feature (Fluidity). Fluidity appears to peak around central values for both *W* ^*S*^ and 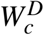, whereas no clear dependence is observed for 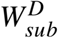, consistent with the fact that Fluidity is a global feature, while subcortical dopaminergic drive primarily affects a limited set of subcortical regions.

Inference validity was assessed through posterior predictive checks (Fig. 5B, bottom right), visualized in a 2D PCA projection of the 72-dimensional data feature space. The small colored points represent the simulations from the SBI training datasets, while red points correspond to data features extracted from simulations generated by sampling the inferred posterior distribution. The clustering of posterior predictive samples near the empirical point (black point) and each other indicates that the SBI framework captured the relationship between parameters and data features, confirming the validity of the inference.

Finally, we evaluated the effect of medication in silico (Fig. 5C). From the posterior distribution of the pathophysiological parameters, we selected the 100 parameter sets whose simulations were closest to the empirical baseline data, defining the unmedicated condition. Starting from these, we simulated 20 increasing doses 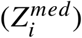 of four antipsychotics (amisulpride, aripiprazole, clozapine, and olanzapine) by increasing or decreasing the pharmacological parameters Z governing receptor occupancy (D_1_, D_2_, and 5-HT_2*A*_), as described in the synthetic experiments. Because post-treatment fMRI data were available, we were able to directly compare simulated medication effects with the empirical post-treatment data. In Fig. 5C, we selected two representative feature pairs to illustrate the key ideas: (i) Fluidity vs. FC between Left Caudate and Hippocampus, and (ii) FC between Right putamen–Insula vs. Right Caudate–Hippocampus. The empirical transition from before and after medication is reflected by a shift in the feature space from the purple to the red star. The simulated medication trajectories move in a similar direction and cluster around the empirical point after medication, indicating that the virtual brain twin captures the observed functional changes induced by treatment. Differences between medications are relatively subtle, likely reflecting overlapping receptor profiles and the limited number of neurotransmitter systems explicitly modeled. The conversion method between the receptor occupancy scaling and the drug dose is explained in the Methods section. We reported the medication effectiveness score for the four antipsychotics across 20 values of the receptor occupancy scaling parameter 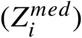 in Fig. 5D, where 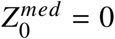 serves as the baseline, corresponding to the absence of pharmacological effect. At each dose level, we performed 100 simulations randomly sampling the pathophysiological paramters 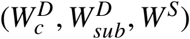 from the inferred parameter posterior distributions. This approach accounts for uncertainty in the inferred pathophysiological parameters, particularly in cases where the posterior distribution remains broad, and avoids biasing the results toward a single estimate such as the mean or the maximum a posteriori. In addition, repeating simulations at each dose reduces the influence of variability in pathophysiological parameters, ensuring that observed changes in functional features are primarily driven by pharmacological manipulation of 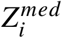. For each simulated dose, we extracted a subset of 9 key functional data features (see Table S3) previously reported to be positively associated with treatment response (*30*). A composite score was then computed by summing each feature that increased relative to the baseline (unmedicated) value. An empirical threshold set at 0.1 was defined to classify treatment effectiveness. The resulting medication effectiveness score are shown for all four medications. In this case, the patient clinically responded to aripiprazole, and accordingly, the simulated response for aripiprazole crosses the effectiveness threshold. A similar trend is observed for clozapine and olanzapine, but not for amisulpride. These results are qualitatively consistent with differential drug efficacy and suggest that the model captures aspects of the patient-specific response to treatment.

#### 2.4.2 Patient 2: Aripiprazole non-responder

We present the results of the virtual brain twin for a patient from the PRONIA dataset who was treated with aripiprazole but did not respond clinically in Fig. 6. Inspection of the posterior distributions reveals differences in the inferred pathophysiological parameters compared to those of Patient 1 described above. In particular, this patient exhibits higher cortical dopaminergic drive and lower subcortical dopaminergic drive, whereas serotonergic drive shows largely overlapping distributions between the two cases.

**Figure 6:**
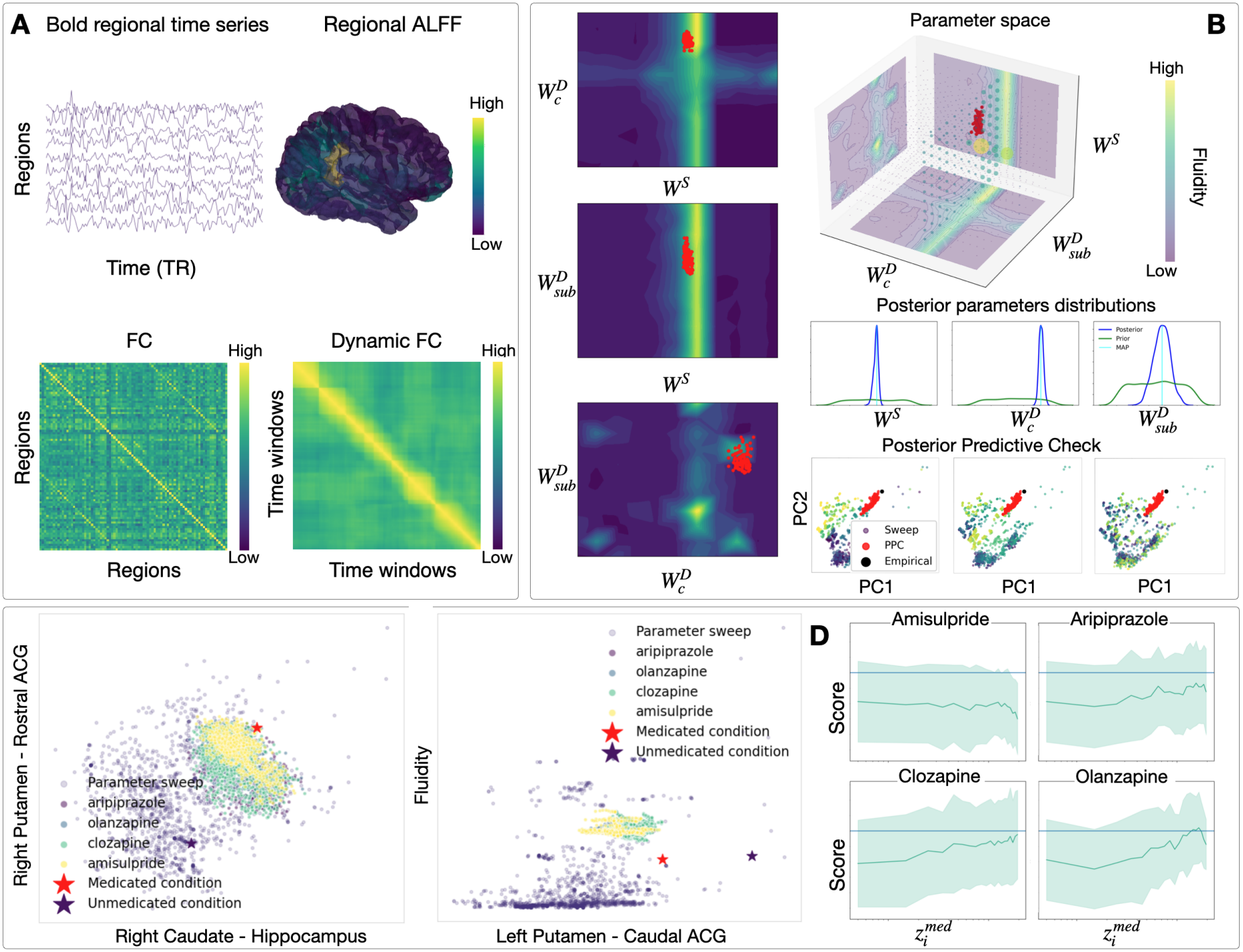
Patient 2: VBT analysis of a non-responder to aripiprazole. Same pipeline and panel organisation as Fig. 5 (Patient 1). (**A**) Empirical BOLD signal and data features. (**B**) Posterior distributions of key pathophysiological parameters inferred by SBI, shown in parameter space and data feature space, ACG = Anterior Cingulate Cortex. (**C**) Medication effects in pairwise data feature space. (**D**) Medication effectiveness scores across increasing doses. The predefined threshold indicates no predicted response for amisulpride, aripiprazole, and clozapine, whereas olanzapine reaches the threshold at higher doses. For this patient, the pipeline would suggest olanzapine as the best treatment option.

The empirical trajectory from baseline to post-treatment and the simulated medication trajectories are shown in two representative data feature spaces: (i) FC between the Right Putamen and Rostral Anterior Cingulate cortex versus FC between the Right Caudate and Hippocampus, and (ii) Fluidity versus FC between the Left Putamen and Caudal Anterior Cingulate cortex (Fig. 6C). In contrast to the responder case, in the dose–response curves (Fig. 6D), aripiprazole does not reach the predefined effectiveness threshold. Similarly, amisulpride and clozapine fail to produce a substantial improvement across the tested dose range. In contrast, olanzapine reaches the threshold at higher values of 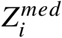, suggesting a potential therapeutic effect in this patient. These findings illustrate how the pipeline could not only predict the failure of aripiprazole but also potentially inform alternative treatment strategies.

#### 2.4.3 Patient 3: Olanzapine responder

We present the results of the virtual brain twin for a third patient from the Xijing Hospital dataseta Xi’an, China. dataset who responded to olanzapine treatment in Fig. 7. Compared with Patient 1, this patient shows lower serotonergic and cortical dopaminergic drive, while subcortical dopaminergic drive shows substantial overlap (Fig. 7B). Relative to Patient 2, this patient exhibits lower serotonergic drive, similar cortical dopaminergic drive, and higher subcortical dopaminergic drive. The empirical trajectory from baseline to post-treatment and the simulated medication trajectories are shown in two representative data feature spaces in Fig. 7C. The medication effectiveness scores indicate that both amisulpride and olanzapine reach the effectiveness threshold, with high doses of aripiprazole also approaching or exceeding this threshold, whereas clozapine does not show a comparable effect (Fig. 7D). Overall, the pipeline would have suggested amisulpride or olanzapine as potential treatment options. Consistent with this prediction, olanzapine was clinically effective for this patient.

**Figure 7:**
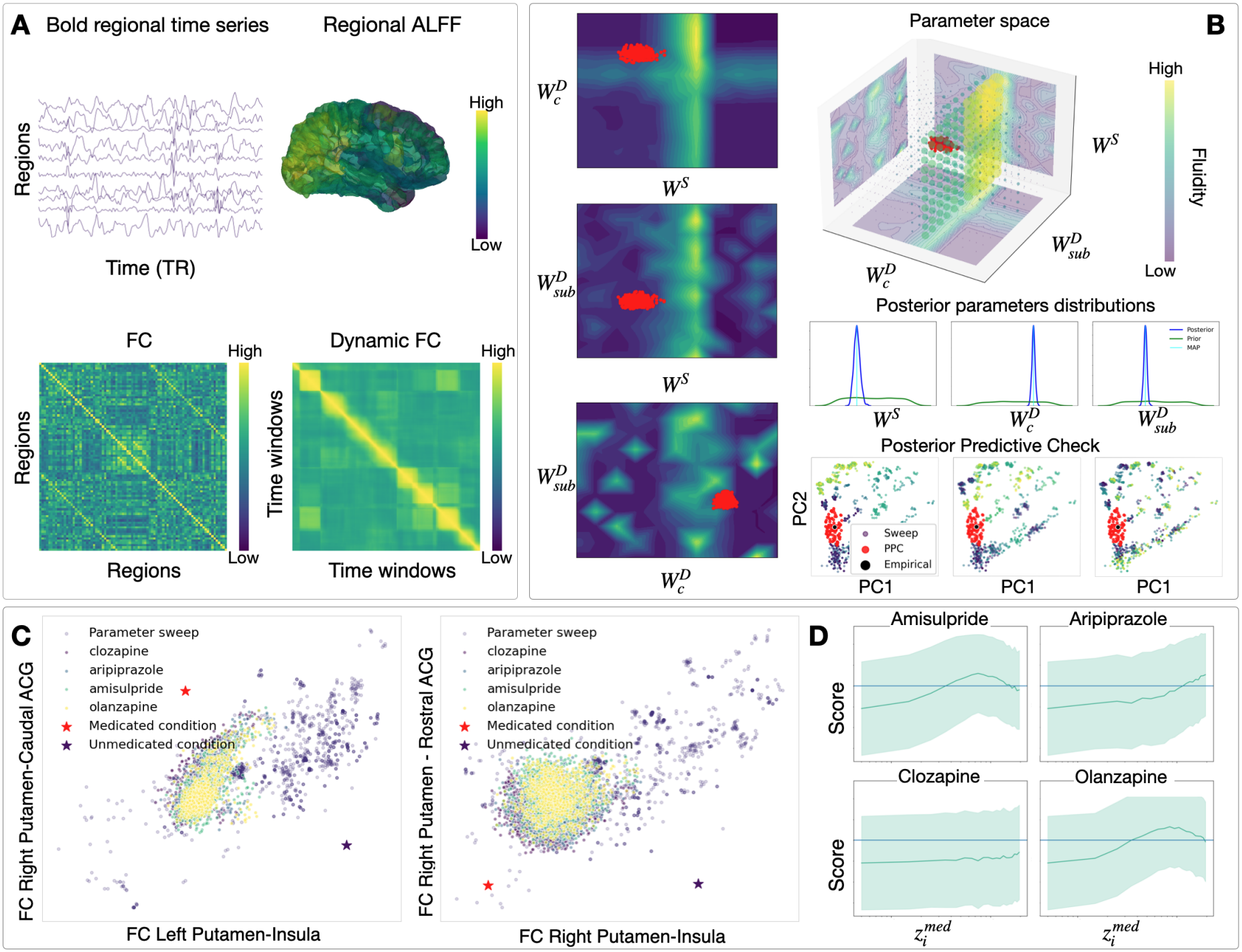
Patient 3: VBT analysis of a treatment responder to olanzapine. Same pipeline and panel organisation as Fig. 5 (Patient 1) and Fig. 6 (Patient 2). (**A**) Empirical BOLD signal and data features. (**B**) Posterior distributions of key pathophysiological parameters inferred by SBI, shown in parameter space and data feature space. (**C**) Medication effects in pairwise data feature space. (**D**)Medication effectiveness scores across increasing doses. The predefined threshold indicates a predicted response for amisulpride, aripiprazole, and olanzapine, whereas clozapine does not reach the threshold.

#### 2.5 Pathophysiological parameters

We analysed differences in the pathophysiological parameters 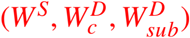 across five comparisons. First, we compared parameter estimates between 9 Healthy Controls (HC) and 9 individuals with First-Episode Schizophrenia (FES) from Shanghai Pudong New Area Mental Health Center (Shanghai, China). In the PRONIA dataset, for which fMRI data were available both at baseline and follow-up for each patient, we assessed differences between 14 Responders and 15 Non-responders at baseline and follow-up, as well as longitudinal changes from baseline to follow-up within each group. To characterize these differences, we performed three complementary analyses. First, we repeatedly sampled one value from each subject’s posterior distribution and computed the corresponding group mean at each iteration. The difference Δ between the sampled group means was then calculated for each iteration, yielding a Monte Carlo distribution of the between-group difference. The 95% credible interval (CI) was defined by the 2.5^th^ and 97.5^th^ percentiles of this distribution, and the probability of a positive between-group difference, *P*(Δ > 0), was estimated as the proportion of sampled differences greater than zero. Second, we compared the overall parameter distributions between groups using the Kolmogorov–Smirnov (KS) distance. For each group, we pooled the individual posterior samples by flattening and concatenating them into a single distribution, and then computed the KS distance between the resulting distributions. Finally, we calculated the mean of each subject’s posterior distribution to obtain a single parameter estimate per subject.

We then compared these subject-level mean estimates between groups using independent-samples, two-sided t-tests. When longitudinally comparing the same subjects at baseline and follow-up, we used a paired t-test.

Figure 8A illustrates the distributions of the sampled group means for HC (yellow) and FES (purple). The between-group difference was defined as Δ = *FE S* − *HC*, and we quantified the probability that this difference was positive, *P*(Δ > 0). For the 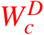 parameter, the 95% CI for the between-group difference was [-1.386, -0.330], with *P*(Δ > 0) < 0.001. This suggests a higher cortical dopaminergic drive in HC than in FES, consistent with theories of cortical hypodopaminergia in schizophrenia. The KS distances between the pooled posterior distributions were 0.13, 0.45, and 0.31 for *W*^*S*^, 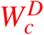, and 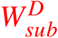, respectively, indicating the greatest distributional separation for cortical dopamine. However, the two-sided t-tests did not report any statistically significant between-group differences for any parameter. Complete results are reported in Table 1. Figure 8B shows the distribution of shrinkage values achieved by SBI for each inferred parameter. Mean shrinkage values close to 1 (also reported in Table 6) indicate a substantial reduction in uncertainty relative to the prior distribution.

**Figure 8:**
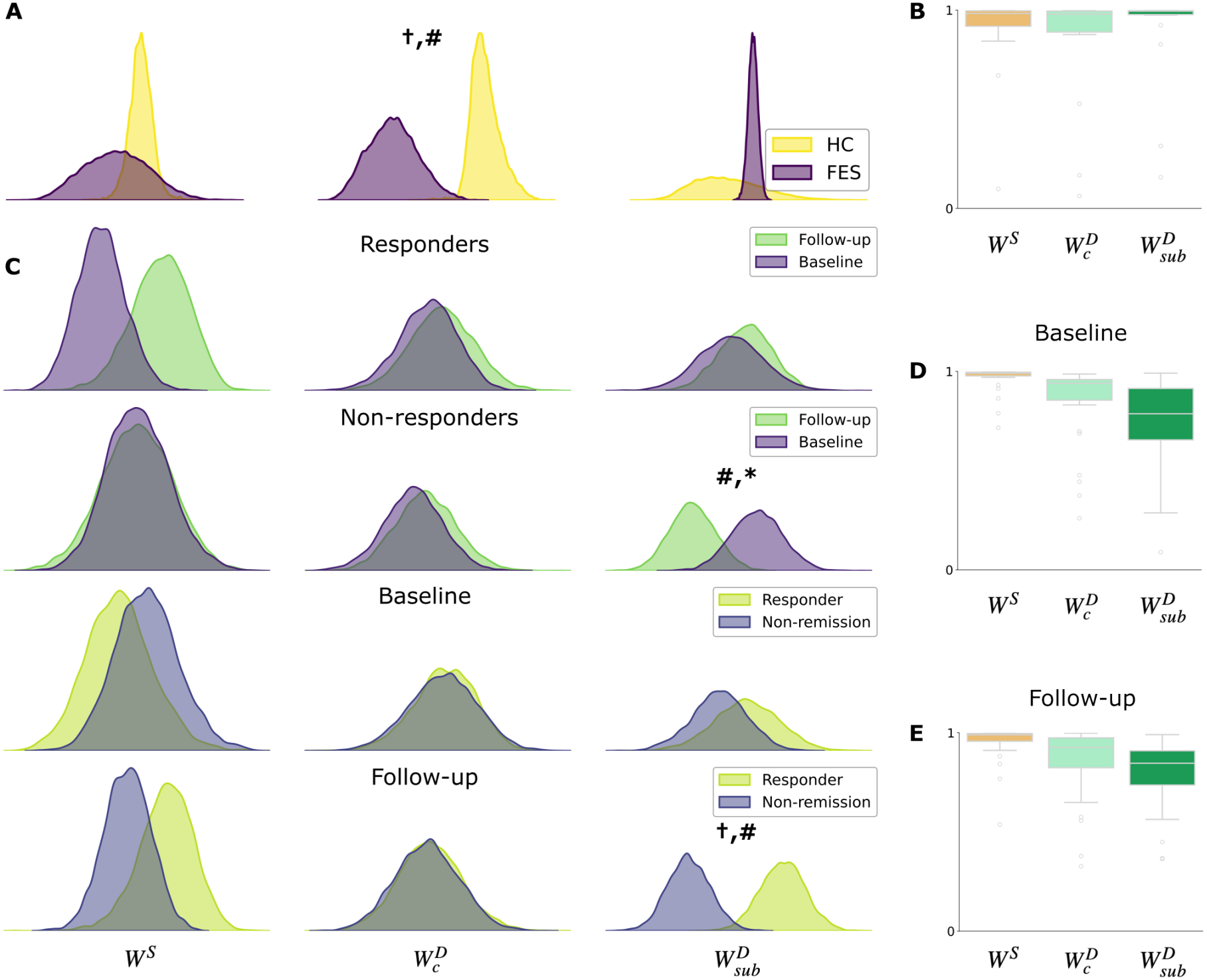
Comparison of the mean posterior distributions of the inferred pathophysiological parameters with corresponding shrinkage across different groups of subjects. (**A**) Distributions of the sampled group means for HC (yellow) and FES (purple). For 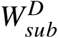, HC show a greater distribution than FES (†, #). (**B**) Distribution of shrinkage values of the inferred parameters of the HC and FES groups. (**C**) Distributions of the sampled group means for the longitudinal changes from baseline (in purple) to follow-up (in green) within each group of Responders and Non-responders, as well as the comparison of the Responders (in yellow) and Non-responders (in blue) at baseline and follow-up. For Non-responders, 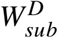 at baseline is lower than at follow-up (#, *). For 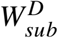 at follow-up, Responders show greater distribution than Non-responders ( †, #). (**D**) Distribution of shrinkage of the parameters inferred at baseline. (**E**) Distribution of shrinkage of the parameters inferred at follow-up.

Figure 8C illustrates the distributions of the sampled group means for the longitudinal changes from baseline (in purple) to follow-up (in green) within each group of Responders and Non-responders, as well as the comparison of the Responders (in yellow) and Non-responders (in blue) at baseline and follow-up. Among Non-responders, 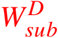 showed a tendency to decrease from baseline to follow-up. Although the 95% CI included zero, the posterior probability of a negative change was high, corresponding to *P*(Δ > 0) = 0.033 for Δ = follow-up − baseline. This pattern was also supported by the paired t-test (*p* ≈ 0.039), although the relatively small Kolmogorov– Smirnov distance suggests limited separation between the pooled baseline and follow-up posterior distributions. Additionally, the mean distribution of 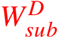 at follow-up could separate Responders from Non-responders (Δ = Responders − Non-Responders). The probability that this difference was positive, *P*(Δ > 0) > 0.999, (95% CI [0.247, 1.114]), suggesting that Responders kept a higher subcortical dopaminergic drive compared to Non-responders. Across all combination of comparisons, this one is also the one with highest KS distance (0.25). However, the independent t-tests did not report a statistically significant between-group differences (*p* ≈ 0.054), and it was statistically not significant for any other comparison neither. Taken together these findings may suggest a potential association between subcortical dopaminergic drive and response to treatment. 8D and E show the distribution of shrinkage values achieved by SBI for each inferred parameter at baseline and follow-up. In both cases, 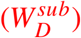 exhibited lower shrinkage than the other parameters, although its mean shrinkage remained relatively high. Full results of the statistical tests are reported in the Tables 2, 3, 4, 5 below.

**Table 2.** Comparison of inferred parameters between follow-up and baseline in responders. The difference is defined as follow-up − baseline.

| **Parameter** | **Mean difference [95% CI]** | *P*(Δ > 0) | **KS distance** | **t-test p-value** |
|---|---|---|---|---|
| *W*<sup>*S*</sup> | 0.154 [−0.041, 0.349] | 0.942 | 0.200 | 0.42 |
| *W*<sub>*c*</sub><sup>*D*</sup> | 0.063 [−0.278, 0.403] | 0.618 | 0.181 | 0.87 |
| *W*<sub>str</sub><sup>*D*</sup> | 0.094 [−0.410, 0.599] | 0.647 | 0.076 | 0.68 |

**Table 3.** Comparison of inferred parameters between follow-up and baseline in non-responders. The difference is defined as follow-up − baseline.

| **Parameter** | **Mean difference [95% CI]** | *P*(Δ > 0) | **KS distance** | **t-test p-value** |
|---|---|---|---|---|
| *W*<sup>*S*</sup> | −0.008 [−0.190, 0.175] | 0.495 | 0.110 | 0.98 |
| *W*<sub>*c*</sub><sup>*D*</sup> | 0.057 [−0.305, 0.418] | 0.606 | 0.135 | 0.84 |
| *W*<sub>str</sub><sup>*D*</sup> | −0.422 [−0.872, 0.029] | 0.033 (#) | 0.155 | 0.039 (*) |

**Table 4.** Comparison of inferred parameters between responders and non-responders at baseline. The difference is defined as Responders − Non-responders.

| **Parameter** | **Mean difference [95% CI]** | *P*(Δ > 0) | **KS distance** | **t-test p-value** |
|---|---|---|---|---|
| *W*<sup>*S*</sup> | −0.050 [−0.223, 0.123] | 0.267 | 0.243 | 0.78 |
| *W*<sub>*c*</sub><sup>*D*</sup> | 0.005 [−0.341, 0.350] | 0.517 | 0.119 | 0.97 |
| *W*<sub>str</sub><sup>*D*</sup> | 0.155 [−0.350, 0.659] | 0.742 | 0.093 | 0.58 |

**Table 5.**
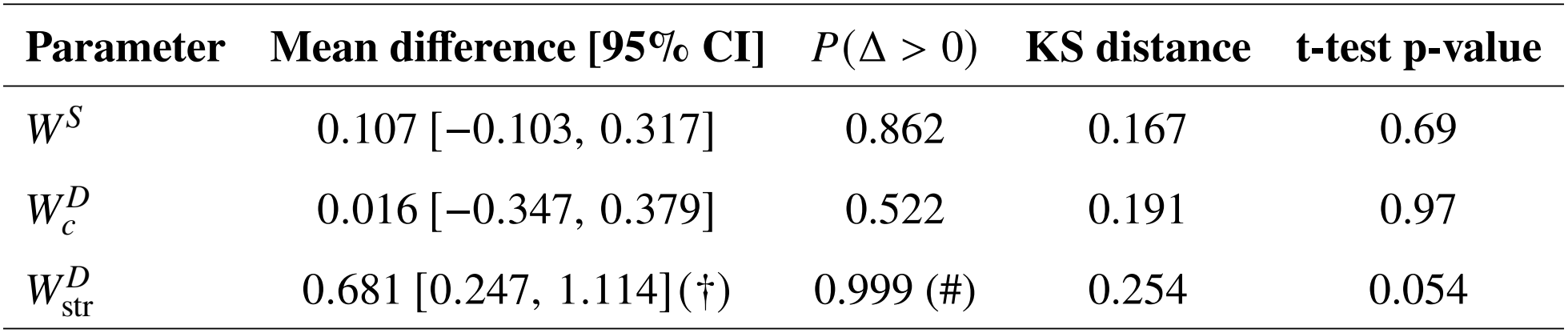
Comparison of inferred parameters between responders and non-responders at follow-up. The difference is defined as Responders − Non-responders.

| **Parameter** | **Mean difference [95% CI]** | *P*(Δ > 0) | **KS distance** | **t-test p-value** |
|---|---|---|---|---|
| *W*<sup>*S*</sup> | 0.107 [−0.103, 0.317] | 0.862 | 0.167 | 0.69 |
| *W*<sub>*c*</sub><sup>*D*</sup> | 0.016 [−0.347, 0.379] | 0.522 | 0.191 | 0.97 |
| *W*<sub>str</sub><sup>*D*</sup> | 0.681 [0.247, 1.114](†) | 0.999 (#) | 0.254 | 0.054 |

**Table 6.** Mean posterior shrinkage for each inferred parameter across the Shanghai-Pudong and the PRONIA cohorts.

| **Comparison** | *W*<sup>*S*</sup> | *W*<sub>*c*</sub><sup>*D*</sup> | *W*<sub>str</sub><sup>*D*</sup> |
|---|---|---|---|
| Shanghai – HC vs FES | 0.90 | 0.85 | 0.89 |
| PRONIA – Baseline | 0.96 | 0.86 | 0.74 |
| PRONIA – Follow-up | 0.95 | 0.86 | 0.79 |

## 2.6 Prediction of medication efficacy

We validate the accuracy of the pipeline in predicting treatment outcomes on 29 patients from the PRONIA dataset who were treatment-naive at baseline, when they were first scanned, and who were treated with one of the antipsychotics from our list (amisulpride, aripiprazole, clozapine, or olanzapine) at their re-evaluation at follow-up (Fig. 9A). Among these patients, 14 met the criteria for remission according to the Andreasen criteria (*31*), while 15 did not. There were four cases of concurrent treatment with more than one antipsychotic at follow-up. For these patients, we only considered the last drug added to the patient’s therapy. This choice was made because the last antipsychotic was added after the patients experienced a new psychotic episode, indicating that the previous treatment had not been effective. In the pipeline, a medication is considered effective if its peak on the dose-response curve reaches a threshold value of 0.1. This value was first optimized to maximize the accuracy of the prediction on the subset of 9 patients who were only treated with aripiprazole during the study period. We then applied this same threshold value to the cohort of 29 patients. Results of the 9 initial patients are available in the Supplementary Fig. S4.

**Figure 9:**
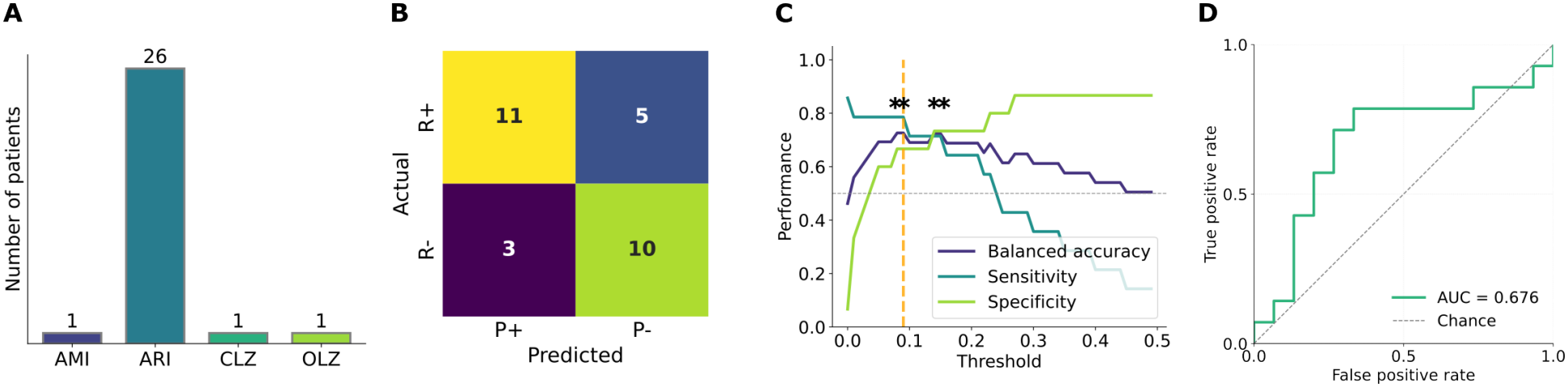
Accuracy of the pipeline prediction. (**A**) Bar plot showing the number of patients receiving each antipsychotic as their last treatment at follow-up. (**B**) Contingency table showing the stratification of the 29 patients according to their empirical treatment response and pipeline prediction. (**C**) Sweep across threshold values showing the corresponding sensitivity, specificity, and balanced accuracy. The orange line indicates the threshold optimized on the smaller dataset and subsequently applied to the larger cohort. Asterisks indicate threshold values for which the two-sided binomial test of accuracy against chance-level performance gives *p* < 0.05. (**D**) ROC curve computed across increasing threshold values.

To assess the predictive performance of the pipeline, we constructed a contingency table (Fig. 9B). Patients were stratified according to their empirical treatment response (responders, R+; non-responders, R-) and the pipeline prediction (predicted responders, P+; predicted non-responders, P-). The results show 11 true positives, 5 false negatives, 3 false positives, and 10 true negatives. The sensitivity of this classification is 78% (Clopper-Pearson confidence interval [49% − 95%]), the specificity is 67% (CI [38% − 88%]), and the balanced accuracy is 72% (CI [44% − 92%]). We performed a binomial test against the null hypothesis that the probability of a correct prediction is 0.5 (i.e., chance-level performance), which gives *p* ≈ 0.024. We then investigated the role of the threshold value by sweeping it to assess how these metrics changed accordingly. Fig. 9C reports the sensitivity, specificity, and balanced accuracy obtained at each threshold. The original threshold used for the prediction displayed in the contingency table is marked in orange, while asterisks indicate the threshold values for which the binomial test gives *p* < 0.05. Using the threshold sweep, we then computed the ROC curve, which shows the relationship between the false positive rate and true positive rate across different threshold values, with an AUC of 0.676.

## 3 Discussion

In this work, we present the first computational pipeline for generating personalized VBT of patients with schizophrenia, with the goal of supporting future clinical decision-making for its treatment. The pipeline constructs subject-specific brain network models capable of simulating both electrophysiological activity and BOLD signals by integrating three subject-specific neuroimaging modalities — T1-weighted MRI for definition of brain regions and cortical thickness estimation, diffusion-weighted imaging for individual connectome reconstruction, and resting-state fMRI for functional feature extraction. Crucially, these simulations reproduce empirical functional features observed in individual patients. The pipeline is further equipped with an SBI framework that enables efficient Bayesian estimation of pathophysiological parameters, and with a pharmacological perturbation module that simulates the effects of antipsychotic medications in silico. To our knowledge, this is the first pipeline to integrate personalised structural neuroimaging, functional inference, and medication simulation within a unified VBT framework for schizophrenia. We applied this pipeline for the first time on three cohorts, investigating the pathophysiology of the disease and reproducing the effects of its pharmacological treatment in silico.

The inferred pathophysiological parameters showed patterns broadly consistent with current neurobiological hypotheses of schizophrenia. Reduced cortical dopaminergic drive in FES patients relative to healthy controls align with the well-established dopamine hypofrontality hypothesis of schizophrenia. Although the small cohort size limits statistical power, the directional consistency of these findings with existing literature lends support to the biological plausibility of the inferred parameters.

At the level of treatment response, remitting patients showed an estimated subcortical dopaminergic drive at follow-up that was higher than non-responders, while non-responders showed a tendency toward a reduction in this drive between baseline and follow-up. These results are compatible with the hypothesis that subcortical dopaminergic dysregulation is relevant to schizophrenia. However, their interpretation in light of the classical subcortical hyperdopaminergia hypothesis is not straightforward. One might expect a reduction in subcortical dopaminergic drive among responders, potentially reflecting a shift toward a more physiological state. Conversely, persistently elevated dopaminergic drive might be expected in non-responders. The observed pattern does not conform to these expectations and highlights the complexity of the relationship between model-inferred dopaminergic drive and clinical response. Several factors may contribute to this discrepancy, including interindividual pathophysiological heterogeneity and differences in treatment regimens. Indeed, most of the patients were treated with aripiprazole, which is known to act as a partial agonist at D2 receptors, increasing dopaminergic signalling under conditions of low endogenous dopamine activity and reducing it when dopamine activity is high. This pharmacological profile could partly contribute to the observed pattern, as its effects may depend on patients’ baseline dopaminergic state. These findings therefore highlight the importance of stratifying patients not only by treatment, but also by their baseline model-inferred subcortical dopaminergic drive. Such analyses could help determine whether the association between dopaminergic drive and treatment response differs according to the pharmacological mechanism of the administered antipsychotic and patients’ initial pathophysiological profiles. These findings should therefore be interpreted cautiously and require further investigation in larger cohorts, with treatment-specific stratification. Nevertheless, the differential parameter trajectories between remitters and non-remitters suggest that the VBT pipeline may capture patient-specific neurochemical dynamics that are relevant to treatment outcome. Future work could integrate these parameters or even related dynamics into treatment response prediction and medication effectiveness score calculation to improve predictive validity.

Several features of the present pipeline distinguish it from existing VBT pipelines developed for other neurological conditions, such as epilepsy (*32*), Parkinson’s (*33*) and multiple sclerosis (*34*). First, subject-specific structural connectomes were annotated by dominant neurotransmitter modality — glutamatergic, GABAergic, dopaminergic, and serotonergic — derived from individual DWI data combined with literature-derived receptor density maps. This integration of personalised white matter architecture with neurotransmitter-specific pathway labelling represents a level of structural personalisation not previously achieved within a unified computational model of brain disorders. Second, the SBI framework enables principled Bayesian inference over a high-dimensional parameter space without requiring explicit likelihood computation, making it both computationally tractable and statistically rigorous. The validity of this inference was confirmed through posterior predictive checks and, in the case of synthetic patients, through direct comparison with known ground truth parameters, confirming that the inferred posteriors accurately recover the underlying neurochemical states and reproduce empirical functional features. Third, the pharmacological perturbation module simulates dose-dependent receptor occupancy effects on neural dynamics, providing a mechanistically grounded and quantitative framework for in silico medication testing. Together, these components constitute a flexible and extensible pipeline that can, in principle, be adapted to other brain disorders, treatment modalities, or imaging protocols.

The present pipeline represents a first foundation toward personalised VBT for psychiatric applications, and as such, it opens a number of promising directions for future works. As a first-of-its-kind pipeline, it is built on a set of principled simplifying assumptions, each of which represents a natural avenue for future investigation. Addressing each of these assumptions may improve predictive performance, though whether they represent a true added value will require careful and dedicated validation. We outline these directions below, each of which we believe deserves a study in its own right. A first natural extension concerns the personalisation of receptor density maps. While the pipeline achieves a high degree of subject-specificity in structural and functional neuroimaging, receptor and transporter density maps for neuromodulators are currently derived from PET data averaged across healthy individuals, rather than from individual subjects. Recent work has shown that cortical receptor densities vary more across individuals than across regions, whereas subcortical densities show the opposite pattern (*35*), highlighting the importance of subject-specific receptor mapping. Furthermore, potential alterations in receptor density associated with schizophrenia due to underlying pathophysiology or long-term pharmacological treatment (*36, 37*) are not currently accounted for. We believe that incorporating individualised receptor distributions represents a critical next step toward better biological realism and may improve parameter identifiability. In the context of a potential clinical application of this pipeline, patient-specific PET scans would be costy and unpractical in clinical routine. However, as subject-specific PET data become more widely available, future work could assess the impact of incorporating such data, for example, in a subset of patients with available PET scans, or use them as ground truth to evaluate the feasibility of inferring receptor densities using an SBI approach.

The second avenue for future work concerns the reconstruction of midbrain projections. The spatial resolution of current DWI methods does not permit precise estimation of fiber tracts connecting midbrain nuclei to cortical and subcortical regions. To partially address this, we evaluated the relationship between D1 receptor density and structural connectivity derived from prior work, observing a strong correlation (≈ 76%), supporting the use of scaled receptor density as a reasonable proxy (*38*). Nevertheless, current tractography methods cannot disentangle the contributions of different neurotransmitter modalities within the same anatomical connection, nor reliably capture directionality. To manage these limitations, the present model constrains each region to a single dominant connection modality and assumes symmetric connectivity weights, while the connectivity mask 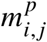 allows partial asymmetry by specifying neurotransmitter type at the node level. Moreover, a robustness analysis (Fig. S3) allowed us to assess the stability of the operating regime of the model under perturbations in the neuromodulatory connectivity structure. Future work could leverage higher-resolution diffusion imaging and directional tractography methods to better resolve midbrain projection pathways and capture the asymmetric nature of neurotransmitter-specific connections, ultimately improving the biological fidelity of the model.

A third direction for future work should investigate more neurotransmitter systems to be included in the model. Other major neurotransmitter systems extensively studied in schizophrenia that were not considered could alter the predictions of the model, limiting its practical value. Additional systems not currently included — such as acetylcholine (*39*), histamine (*40*), and further serotonergic receptor subtypes (*41*) — represent promising targets for future model extension. The present framework deliberately focuses on dopamine (D_1_/D_2_) and serotonin (5-HT_2_ *A*), although not explicitly representing presynaptic D_2_ autoreceptor-mediated regulation of dopamine synthesis and release, which may influence the effects of dopaminergic modulation. Nevertheless, these systems have well-established roles in schizophrenia pathophysiology and antipsychotic pharma-cology, allowing tractable parameter estimation while capturing the core neurochemical drivers of the disorder. Establishing this foundation first allows each subsequent extension to be evaluated against a well-characterized baseline, so that the specific contribution of each additional system can be assessed rigorously. Incorporating these systems would increase biological realism but would also substantially raise computational complexity and the risk of parameter degeneracy, whereby different biological mechanisms produce indistinguishable model outputs. This risk is inherent to the current model architecture, in which neural dynamics are represented through excitatory (AMPA), inhibitory (GABA), and leakage currents, constraining the parameter space onto which multiple neurotransmitter effects must be mapped. Expanding the neurotransmitter repertoire is therefore a promising but technically demanding direction that must be balanced carefully against the need for parameter identifiability and computational tractability.

A further direction concerns the model’s representation of cortical pathology. In the present framework, cortical thickness derived from T1-weighted MRI is used to inform regional excitability parameters, providing an indirect link to glutamatergic function. This choice is motivated by converging evidence that cortical grey matter volume loss and decreased glutamatergic activity, both consistently reported in schizophrenia, may reflect a common underlying cellular mechanism (*17, 21, 22*). Post-mortem studies support this interpretation, demonstrating consistent reductions in the size of dendrites, axons, and cell bodies of cortical glutamatergic neurons, as well as decreased synaptophysin expression, a marker of presynaptic axon bouton density, in schizophrenia (*23*). Taken together, these findings suggest that cortical volume loss may serve, at least in part, as a macroscopic correlate of glutamatergic synaptic deficiency, lending biological plausibility to the use of cortical thickness as a proxy for excitatory drive. However, linking macroscopic volume loss to specific cellular alterations remains challenging, as volume reduction may reflect synaptic changes, variations in myelination, neuronal soma size, dendritic modifications, or combinations thereof (*21*). Subcortical gray matter volume is not incorporated in the present framework; consequently, excitatory parameters for all subcortical regions are fixed to default values. Future work should extend the model to account for regional variability in subcortical gray matter volume. Furthermore, GABAergic interneuron populations, which show reduced density in schizophrenia (*42*), are not explicitly parametrised in the present model, representing a limitation that future iterations should address. Incorporating GABAergic dynamics more explicitly would improve the model’s ability to capture the excitatory-inhibitory imbalance increasingly implicated in the pathophysiology of schizophrenia.

A fourth avenue for future work concerns the incorporation of task-based neuroimaging data. Resting-state fMRI provides a rich and clinically accessible source of functional features, and the present pipeline demonstrates that these alone is able to infer meaningful pathophysiological parameters and simulate pharmacological effects in silico. A substantial body of literature has demonstrated that schizophrenia is also characterized by altered brain responses during task-based paradigms, with distinct patterns observed across different stages of the illness, including early psychosis and chronic schizophrenia (*43*). Moreover, task-related activity can be modulated by antipsychotic treatment (*44*). For example, abnormalities in task-evoked responses have been consistently reported in paradigms such as the auditory oddball task (*45, 46*). Incorporating task-based data therefore represents a promising direction for future work, as it provides a more direct link between brain dynamics and behavior.

Finally, future work should aim to incorporate the full trajectory of the disease. This includes not only different clinical stages such as individuals at risk, first-episode patients, and chronic cases, but also the underlying developmental processes. In particular, modeling structural and functional brain changes across childhood and early adolescence could help predict disease onset, progression, and relapse (*47*). Integrating such longitudinal and developmental perspectives would significantly improve the predictive power and clinical relevance of VBT frameworks.

In summary, the work demonstrates that personalised VBT can be constructed from routine clinical neuroimaging data, and that the resulting models capture neurochemically meaningful individual differences relevant to both diagnosis and treatment response. While the pipeline is currently at an early stage of validation, the results support its potential as a tool for in silico treatment planning, enabling clinicians to simulate the likely effects of different medications for a given patient before initiating pharmacological intervention. Crucially, the proposed pipeline will be applied to two prospective clinical trials conducted in Marseille and Munich as part of the Virtual Brain Twin project, funded by the European Union’s Horizon Europe Programme, with data collection scheduled to begin in the second half of 2026. These trials will provide the larger and more diverse cohorts necessary for rigorous prospective validation and will represent a critical step toward establishing VBT-guided treatment selection as a clinically viable approach. The present work provides a principled, validated, and extensible foundation for this translational agenda and a concrete first step toward the clinical application of personalised brain simulation in psychiatry.

## 4 Materials and Methods

### 4.1 Patients Data and Preprocessing

A total of 51 subjects were included in this study. The first dataset consisted of 9 healthy controls and 9 patients with First Episode Schizophrenia (FES) recruited at Shanghai Pudong New Area Mental Health Center, Shanghai, China. This cohort was used to evaluate the model and test the pathophysiological assumptions underlying the simulations (*3*). A second dataset included FES patients recruited from the Department of Psychiatry, Xijing Hospital in Xi’an, China. This dataset included four patients and two healthy controls, each with baseline and follow-up scans after pharmacological treatment, which was used to investigate the effects of simulated in silico drug interventions (*48*). The subjects were aged between 19 and 31 years old, including two male and two female patients, and one male and one female healthy control. Patients were treated with either amisulpride, olanzapine, paliperidone, or risperidone. The patients’ clinical status was assessed using the positive, negative, and general subscales of the PANSS (*49*). Overall, PANSS scores decreased across all dimensions in most patients. However, one patient showed an increase in the positive subscale, and another showed an increase in the general subscale. The third dataset comprises nine patients from LMU Munich Hospital, scanned twice: once prior to treatment (drug-naïve) and once after treatment with aripiprazole. This cohort enabled evaluation of model predictions against longitudinal pharmacological effects. In this cohort, patients were aged between 20 and 40 years old, including eight males and one female. Remission after treatment was evaluated using the Andreasen remission criterion (*31*), according to which 5 achieved remission and 4 did not. All participants underwent comprehensive psychiatric evaluation as well as high-resolution 3T MRI acquisition, including T1-weighted structural imaging, diffusion-weighted imaging (DWI), and resting-state BOLD fMRI. An additional 20 patients from the PRONIA dataset were then added to assess the predictive power of the pipeline. These patients were treated with amisulpride, aripiprazole, clozapine or olanzapine. 6 were females, 14 were males, 10 responded to treatemnt, 10 did not. All demographics are reported in Table 7.

**Table 7.** Summary of the study cohorts.

| **Site** | **Group** | **N** | **Sex (M/F)** | **Age (years)** | **Remission** |
|---|---|---|---|---|---|
| Shanghai Pudong | HC | 9 | 2/7 | 19–29 | – |
| Shanghai Pudong | FES | 9 | 4/5 | 18–37 | – |
| Xijing Hospital, Xi’an | HC | 2 | 1/1 | 19–31<sup>*a*</sup> | – |
| Xijing Hospital, Xi’an | FES | 4 | 2/2 | 19–31<sup>*a*</sup> | 2/4 |
| PRONIA | FES | 29 | 22/7 | 20–40 | 14/29 |
| Total | – | 53 | 31/22 | 18-40 | 16/33 |

HC, healthy controls; FES, first-episode schizophrenia; M, male; F, female. Response in the PRONIA cohort cohort was assessed using the Andreasen remission criteria.
<sup>*a*</sup> The reported age range of 19–31 years was provided for the Xijing Hospital cohort as a whole.

**Table 8:** Normalized receptors affinities of four antipsychotics. Amisulpride is known to have a much higher affinity for D_2_R compared to other receptors, while Aripiprazole and Clozapine predominantly affect 5-HT_2*A*_R, whereas Olanzapine has a similar effect on both D_2_R and 5-HT_2*A*_R. For each drug, normalization is achieved by dividing by the highest affinity.

|  | *Amisulpride* | *Aripiprazole* | *Clozapine* | *Olanzapine* |
|---|---|---|---|---|
| D<sub>1</sub>R | 10<sup>−4</sup> | 0.004 | 0.034 | 0.02 |
| D<sub>2</sub>R | 1 | 0.26 | 0.02 | 0.88 |
| 5-HT<sub>2*A*</sub>R | 10<sup>−4</sup> | 1 | 1 | 1 |

Because the datasets originated from different sites, the imaging protocols differed in acquisition parameters and overall image quality. All statistical analyses and parameter comparisons were performed separately for each dataset, without pooling patients across datasets.

Structural T1-weighted images were processed using the recon-all pipeline of FreeSurfer (*50*), which performs volumetric segmentation and cortical surface reconstruction. The reconstructed cortical surface was parcellated according to the Desikan–Killiany (DK) atlas (*51*). Diffusion MRI data were processed using the MRtrix3 software package (*52*). Whole-brain tractography was performed by generating 5 million streamlines per subject. Structural connectivity matrices were constructed by assigning streamlines to pairs of DK regions and counting the number of streamlines connecting each pair. This procedure yielded an 84 × 84 structural connectivity matrix for each subject. Because tractography does not provide directionality information, the matrices were symmetric. Diagonal elements were set to zero to exclude self-connections within regions. Finally, each matrix was normalized by its maximum value, resulting in connectivity weights ranging between 0 and 1.

Resting-state fMRI data were preprocessed using the fMRIPrep pipeline (*28*). Due to differences in acquisition quality across cohorts, slightly different nuisance regression strategies were applied. For the FES and LMU cohorts, confound regression included the first five aCompCor components, principal components derived from white matter and cerebrospinal fluid signals to capture physiological noise (*29*). For patients at Xijing Hospital, confound regression included signals from cerebrospinal fluid (CSF), white matter (WM), and six motion parameters.

After preprocessing and confound regression, the BOLD time series were band-pass filtered and parcellated using the Nilearn NiftiLabelMasker utilities (*53*). From the cleaned regional time series, we computed several functional metrics using Python (NumPy and scikit-learn): ALFF (Amplitude of Low-Frequency Fluctuations) to quantify regional spontaneous activity, Functional Connectivity (FC) based on pairwise correlations between regional signals, Functional Connectivity Dynamics (FCD) to characterize time-varying connectivity patterns. These measures were used to evaluate the correspondence between empirical data and simulated brain dynamics.

### 4.2 Mean field models with dopaminergic and serotonergic modulation

Brain activity is simulated at each node using a set of differential equations that describe the mean-field population activity of neurons at that node. The model is an adaptation of the work by (*54*), where it was derived from a population of Izhikievic Neurons (*55*), assuming an infinite neuronal population and Lorentzian distributed additive currents centered at *η* with a half-width Δ accountable for the internal variability of the neural mass.

We added the effect of two neuromodulators, dopamine and serotonin, to the original model, following the approach described by (*56, 57*). This modification aims to capture the influence of the neuromodulators on neural excitability, dopamine via the D_1_ and D_2_ type receptors (D_1_ and D_2_R), and serotonin via the 5-HT_2*A*_ receptor (5-HT_2*A*_R), respectively. In cell biology, D_1_ are G-protein-coupled receptors of the Gs type. When activated by their ligand (dopamine), they stimulate the synthesis of cyclic adenosine monophosphate (cAMP), which activates protein kinase A (PKA). PKA, in turn, activates various second messengers, leading to an increase in the excitatory postsynaptic currents (EPSC) of AMPA channels, as well as their synthesis and membrane expression (*58*). On the other hand, D_2_R couple to Gi/o proteins, inhibiting adenylyl cyclase, acting to reduce the excitability of striatopallidal neurons and their response to glutamatergic synaptic input. Though through a different intracellular mechanism, also 5-HT_2*A*_R activation increases pyramidal cell excitability (*59*) by increasing the EPSC, an effect that is antagonized by AMPA receptor antagonists (*60*). For these reasons, in our framework, the effect of dopamine and serotonin acts on the excitatory currents of the mean-field model.

The mean-field model with the implementation of dopamine and serotonin neuromodulation is the following:

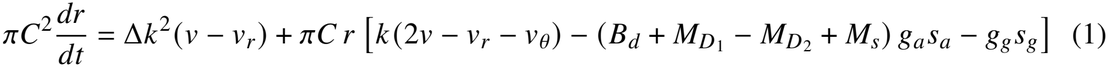

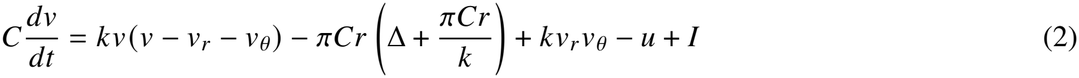

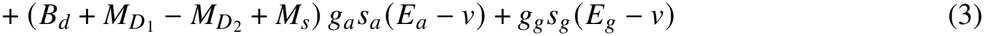

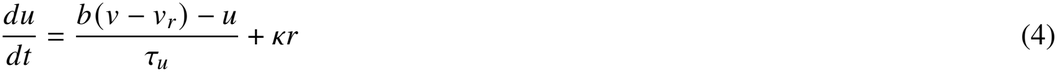

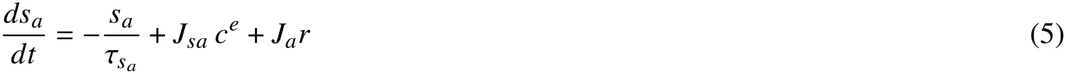

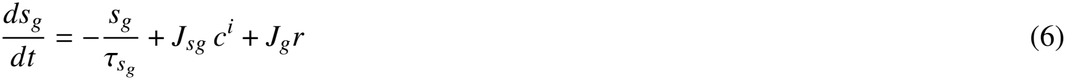

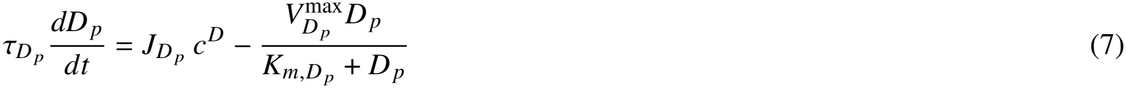

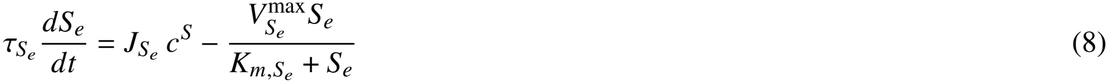

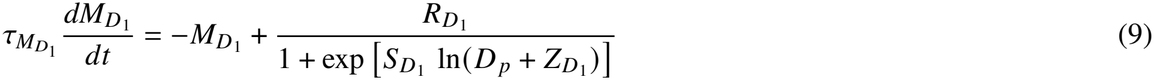

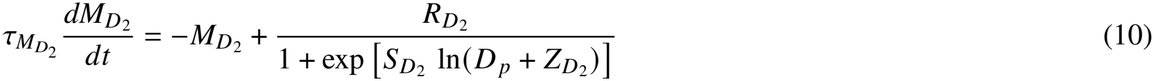

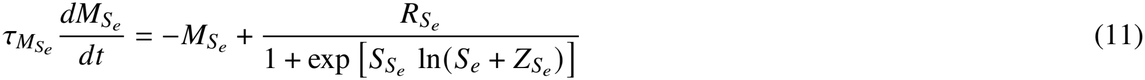

The variables correspond to the firing rate *r*, the mean membrane potential *V*, the adaptation current *u*, the excitatory and inhibitory inputs *s*_*a*_ and *s*_*g*_, the concentration of the neuromodulators *D*_*p*_ and *S*_*e*_ (for dopamine and serotonin respectively), and their neuromodulatory effects 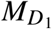, 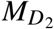, and 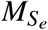. Due to the relatively slow timescale of their concentration changes, the concentration and neuromodulatory variables have significantly higher time constants compared to the others. Note that 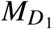 and 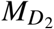 have opposite signs, denoting their opposite effects on the excitatory inputs. Excitatory synaptic currents are mediated by AMPA and NMDA receptors, collectively characterised by a maximal conductance *g*_*a*_ and reversal potential *E*_*a*_, while inhibitory synaptic currents are mediated by GABA receptors of maximal conductance *g*_*g*_ and reversal potential *E*_*g*_. The excitatory synaptic input of a node, *S*_*a*_, depends on the signal received from all the other nodes connected to it, *c*_*e*_, scaled by a factor *J*_*sa*_. Moreover, it accounts for the node’s self-excitatory input *r* scaled by *J*_*a*_. This is the same for the inhibitory input *S*_*g*_. The effect of the neuromodulation is embedded by the multiplicative term 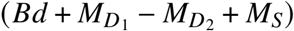 that affects the excitatory current of the node, where *B*_*d*_ is the baseline accounting for synapses in the absence of any neuromodulation. The *M* terms are modeled with a sigmoidal dose-response curve dependent on the neurotransmitter concentration at the node, where *R* are the number of receptors of the neuromodulators and *S* and *Z* are parameters related to the receptor’s affinity and occupancy. The average concentration of any neuromodulator at the node depends on its inputs *c* received by the node, scaled by a factor *J*, and on the reuptake of the neurotransmitter. This latter is described by the Michaelis-Menten equation for the Dopamine Transporter (DAT) and Serotonin Transporter (SERT), which reuptake dopamine and serotonin from the synaptic cleft at a maximal rate *V*^max^ and with a dissociation constant *K*_*m*_. While *R* is extracted from PET data taken from (*61*) and (*62*), *S* and *Z* are rescaled values taken from the work by (*63*). Full parameterization of the equations is shown in Table S1.

### 4.3 Model Implementation and Hemodynamic Modeling

The model was implemented using vbjax, a JAX-based library designed for Virtual Brain–style simulations (*64*). The system of equations was numerically integrated using the Heun method. The firing rate of each brain region, as defined in Equation 1, was used as input to a hemodynamic model to generate the corresponding BOLD signal. Specifically, we employed the Balloon–Windkessel model (*65*), which links neuronal activity to vascular and hemodynamic responses underlying the BOLD signal.

### 4.4 Specific Connectivity

In a model of a full brain, several neural masses are connected to create a network where every node is a neural mass that represents the activity of a specific brain region. The cortical regions are parcellated and segmented from the T1 MRI scans of a human subject, while the links are weighted by the tract numbers extracted from the DW images. In addition, we include six extra segments that will correspond to the neuromodulatory nodes of the network: the Ventral Tegmental Area (VTA) and the Substantia Nigra (SN) for dopamine, the Raphe Nuclei (RN) for serotonin. The nodes need to be connected with some rules. The model allows for four different types of connections: glutamatergic, GABAergic, dopaminergic, and serotonergic. The inputs received by a node can change the activity of the node’s glutamatergic or GABAergic synapses or its dopamine concentration via the *c*^*e*^, *c*^*i*^, *c*^*D*^, and *c*^*S*^ terms, respectively. In general

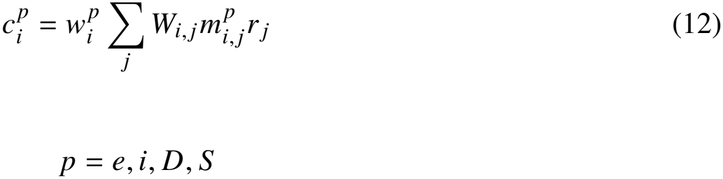

where p is a specific type of projection (glutamatergic, GABAergic, dopaminergic, or serotonergic), and 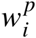 is a scaling factor. *W*_*i, j*_ is the weight of the connection between node i and node j taken from the matrix of the weights of the connectome, and 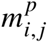 is a value that can be either 0 or 1 depending on the type of projection between nodes i and j taken from the so-called “connectivity masks”, and *r* _*j*_ is the input firing rate from node j. The weights of the connectome *W*_*i, j*_ are obtained from the tractography of the Diffusion MRI Imaging scans of the subject. Since with the current imaging resolution it is difficult to get a precise measure of the weights of the dopaminergic and serotonergic nuclei connections, we manually added to the connectome using the scaled values of the relative receptor density of an area extracted from the PET atlases of (*61*) and (*62*).

Connectivity masks are boolean matrices specific to the type of connection (glutamatergic, GABAergic, dopaminergic, or serotonergic). For a given connection type, 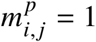 if nodes i and j are connected via that connection type; otherwise, 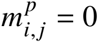. Multiplying the tractography matrix by the corresponding mask isolates the inputs that a node receives from connections of a specific type, ensuring that the node processes only inputs associated with the designated connectivity pattern. Because tractography cannot determine the nature nor the directionality of a connection, the masks must be constructed using anatomical knowledge. To address this, we created a set of connectivity masks based on established anatomical pathways, including: 1) the classical GABAergic circuitry of the basal ganglia, excluding the subthalamic nucleus, which was not segmented (*66*), 2) GABAergic projections from the basal ganglia to both the VTA and SN (*66, 67*), 3) Dopaminergic projections from the VTA to the cortex and from the SN to the basal ganglia (*67–69*), and 4) Serotonergic projections from the RN to the rest of the brain (*70, 71*).

The manual setting of weight and directionality of these projections directly shapes the SBI-inferred posteriors. To investigate the robustness of the model to variations in the connectivity structure, we performed an additional parameter sweep in which we introduced perturbations to the dopaminergic and serotonergic connections (see Fig. S3 of the Supplementary Materials). This analysis allowed us to assess the consistency of the features extracted from the simulations under increasing changes in the imposed connectivity weights, and to verify that the model does not operate in a region of parameter space where even minor perturbations could lead to substantial changes in its dynamics, thus biasing the SBI results.

### 4.5 Grey matter modeling

Assuming glutamate is the neurotransmitter involved, in this work we focus on parameters related to AMPA currents and evaluate how they might link to cortical thickness. Specifically, we introduce regional variability in terms of different cortical thickness of the brain areas by varying two key parameters The first, *J*_*a*_ (local connectivity), describes the total connection strength between neurons within the same node in the mean-field model. Its behavior depends on what it represents: if tied to the number of neurons, *J*_*a*_ decreases as neuron count decreases. If tied to neuronal density (connections per unit volume), *J*_*a*_ remains constant despite neuron loss. If the neuropil volume is reduced, Ja decreases, indicating weaker self-excitation within the node. The second, *J*_*sa*_ (Scaling of Glutamatergic Connections Between Nodes), reflects the strength of connections between neurons received from the other nodes, summed across the population. Similar to *J*_*a*_, its changes depend on its interpretation: if tied to the number of neurons, *J*_*sa*_ decreases with fewer neurons; if tied to neuronal density, *J*_*sa*_ remains unchanged; if the neuropil volume decreases, *J*_*sa*_ decreases, signaling weaker inter-node glutamatergic input.

Cortical gray matter thickness for each cortical brain region was extracted from the statistical output files generated by the FreeSurfer preprocessing pipeline. These regional measurements were then compared against a normative reference model obtained from the CENTILE framework (*24*), which provides population-based estimates of expected cortical thickness across the lifespan. Since normative cortical thickness scores are expressed as z-scores (mean 0, standard deviation 1), we linearly rescaled them to obtain model parameters with mean 12 and standard deviation 1.2. The transformation is defined as

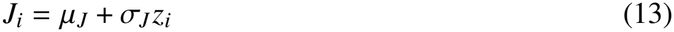

where *z*_*i*_ is the normative cortical thickness z-score for region *i, µ*_*J*_ = 12 is the target mean parameter value, and *σ*_*J*_ = 1.2 is the target standard deviation. These values were chosen after an optimization process that aimed at maximizing the correlation between the empirical and simulated ALFF of cortical regions, with simulations run combining different *µ*_*J*_ and *σ*_*J*_ (see S1).

All the *J* parameters of subcortical regions were set to 12.5, and regions in the midbrain for which it was set to 17.5, in order to mimic a pacemaker-like activity (*72*).

### 4.6 Medication effects

In this model, the neuromodulatory variables *M* are represented by a sigmoidal function that captures a dose-response relationship between a ligand and its receptor (Eq. 9-11). This formulation allows the simulation of pharmacological interventions. Most antipsychotic drugs act as reversible, competitive antagonists at dopaminergic D_1_ and D_2_ receptors, or serotonergic 5-HT_2*A*_ receptors. “Reversible” indicates that the drug does not permanently bind to the receptor, leaving the number of available receptors unchanged. “Competitive” means that the drug competes with the endogenous neurotransmitter (dopamine or serotonin) for the same binding site, thereby reducing the receptor’s effective affinity for its ligand. As a consequence, higher concentrations of neurotransmitter are required to achieve the same receptor activation when antagonist levels increase (*25*).

In the model, the influence of a reversible, competitive antagonist is captured by decreasing the parameter *Z* in the sigmoidal function. As the concentration of the antagonist increases, its receptor occupancy increases as well, and this reflects in a change of *Z*. Lower *Z* values make the neuromodulatory variable less sensitive to the neurotransmitter concentration, effectively reducing receptor activation. Conversely, higher *Z* values simulate the effect of an agonist, increasing the sensitivity of *M* to the ligand.

Using this framework, we simulated the effects of four antipsychotic drugs: clozapine, amisul-pride, olanzapine, and aripiprazole, on D_1_R, D_2_R, and 5-HT_2*A*_R. While these drugs also interact with other receptor types, explicitly modeling all receptor interactions is complex, so we focused on these three primary mechanisms. For each drug, we constructed a mechanism-of-action profile that quantifies its proportionate effect on each receptor based on the normalized receptor affinity data (*73–75*), summarized in Table 8. The baseline values of the *Z* parameters for each receptor are adapted from (*63*). For each medication, we identify the receptor for which it exhibits the highest affinity. Starting from the baseline value of this receptor, we gradually reduce its *Z* parameter towards zero. Thus, the baseline value represents the absence of medication effects, whereas zero represents the maximum antagonist effect. The Z parameters of the other two receptors, for which the medication has lower affinity, are only partially reduced, proportionally to their affinity values normalized by the one of the receptor with the highest affinity. In this way, the largest effect is assigned to the receptor for which the medication has the highest affinity, while smaller effects are assigned to the other two receptors. The rescaling formula of *Z* for a drug at increasing doses is shown in Equation S1. The dose levels explored in the model can be expressed in clinically interpretable units of mg/day, facilitating the interpretation of the simulated treatment trajectories. The conversion is done using a pharmacokinetic–pharmacodynamic framework described by (*27*), which takes into account the molecular weight of each medication, its receptor affinity (Ki), the desired receptor occupancy, the dose-to-receptor concentration conversion factor, and the unbound plasma fraction. The conversion is formulated by Eq. S2.

Aripiprazole act as a partial agonist on D2 receptors (*76*). Its presence increases dopaminergic signalling under conditions of low endogenous dopamine activity and reducing it when dopamine activity is high. To reproduce this effect when simulating aripiprazole administration, its dose-response curve for *M*_*D*2_ (Eq. 10) is shifted to the right and its plateau is lowered. This is achieved by decreasing *R*_*D*2_, which determines the plateau of the curve, and gradually increasing *Z*_*D*2_.

The medication effectiveness score was computed by comparing simulated functional features under pharmacological modulation with their corresponding baseline values obtained in the absence of medication. The feature set included eight functional connectivity measures between two given regions and one dynamic metric: Fluidity (see S3 for details). The total medication effectiveness score is obtained by summing the change in the values of the features when the medication is applied. This scoring approach was motivated by prior evidence indicating that the selected functional connectivity measures are positively associated with clinical response to antipsychotic treatment (*30*). In addition, Fluidity was included based on findings that higher dynamical variability is characteristic of healthy brain function, particularly in younger individuals (*7, 77*). Under this assumption, increases in Fluidity were interpreted as reflecting a shift toward a healthier dynamical regime.

### 4.7 Individualized and population-derived data in the pipeline

The workflow integrates both subject-specific and population-derived data. Subject-specific information includes T1w structural MRI, DWI) and fMRI, from which individualized features are extracted, including brain parcellation, cortical thickness, structural connectomes, and functional data features. Population-derived information consists of the PET atlas from which receptor density maps are obtained and used as proxies for serotonergic and dopaminergic projections (Eq. 12), as well as the receptor count R (Eq. 9-11). The pipeline outputs include individualized parameter estimates (*W*^*S*^, 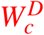, and 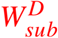) and individualized predictions of treatment response with corresponding effective dose 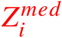.

## Data Availability

All data produced in the present study are available upon reasonable request to the authors

## Acknowledgments

We acknowledge the contribution of the following colleagues and their respective institutions of the PRONIA consortium:

- Lana Kambeitz-Ilankovic, Stephan Ruhrmann, and Joseph Kambeitz — University of Cologne, Germany.
- Georg Romer — University of Münster, Germany.
- Anita Riecher-Rössler — University of Basel, Switzerland.
- Peter Falkai — Medical Centre of Ludwig-Maximilian-University, Munich, Germany.
- Jarmo Hietala and Raimo K. R. Salokangas — University of Turku, Finland.
- Rebekka Lencer — University of Lübeck, Germany.
- Alessandro Bertolino — University of Bari, Italy.
- Paolo Brambilla — University of Milan, Italy.
- Stephen J. Wood — Orygen, National Centre of Excellence for Youth Mental Health, Australia.
- Rachel Upthegrove — University of Oxford, United Kingdom.
- Eva Meisenzahl — University of Düsseldorf, Germany.

## Funding

This project/research has received funding from the European Union’s Horizon Europe Programme under the Specific Grant Agreement No. 101137289 (Virtual Brain Twin Project, to V.J.) and Grant Agreement No. 101147319 (EBRAINS 2.0 Project to V.J). This work has benefited from a government grant managed by the Agence Nationale de la Recherche (ANR) under the France 2030 program, reference ANR-22-PESN-0012 (V.J.).

## Author contributions

Conceptualization: H.W.,N.K., V.J. Methodology: G.P., H.W.,P.S.,D.D. V.J., Formal analysis: G.P., A.Z., Software: G.P., A.Z, M.W.,P.P., P.T., G.C., A.E.,L.D., Validation: A.Z., E.D. M.H.,, Data curation: X.C., M.S., M.F., L.C., J.F.,N.K., Supervision: H.W., V.J. Investigation: V.J., Writing—original draft: G.P. and H.W. Writing—review and editing: All co-authors.

## Competing interests

There are no competing interests to declare.

## Data and materials availability

All data and code needed to evaluate and reproduce the results in the paper are present in the paper and/or the Supplementary Materials. The software code for this study is available at https://github.com/giapre/vbt-project.git.

Supplementary Materials and Methods Figs. S1 to S3

Tables S1

## Supplementary Materials for

### Materials and Methods

#### Parameterization of *J*_*a*_ **and** *J*_*sa*_

The mean and standard deviation of *J*_*a*_ and *J*_*sa*_ were optimized by maximizing the correlation between the empirical ALFF, computed for each cortical region from the BOLD signals of the group of 18 subjects (9 healthy controls and 9 patients with first episode schizophrenia) from the Shanghai Pudong dataset, and the simulated ALFF obtained from simulations run with different values of *µ*_*J*_, *σ*_*J*_, and excitatory coupling *w*^*e*^. This optimization does not yield an individualized optimal parameter set for each patient. Instead, we selected the group-level optimal values of *µ*_*J*_ and *σ*_*J*_, which are shared across all subjects. Subject-specific variability in *J*_*a*_ and *J*_*sa*_ is subsequently introduced based on the regional normative cortical thickness z-scores, as reported in the Results. *J*_*a*_ and *J*_*sa*_ always share the same value. The results of the optimization process are shown in Figure S1.

#### Optimization of excitatory coupling and noise

In the simulations, default values of excitatory coupling (*W* ^*e*^ = 0.3) and noise amplitude (*σ* = 0.15) were used. These values were selected based on a parameter sweep exploring the joint effects of these two parameters on the synthetic patient. For each simulation in the sweep, Fluidity and Global Brain Coupling (GBC) were computed. The explored parameter ranges were chosen to identify regimes that yield physiologically plausible GBC values while maximizing Fluidity. The results of this sweep are shown in Fig. S2, where Fluidity and GBC are represented as heatmaps with *W* ^*e*^ on the x-axis and *σ* on the y-axis.

#### Calibration of other relevant parameters

All the parameters that are not estimated with SBI patient-specifically (*W* ^*s*^,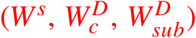), changed under medication effects 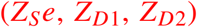, or optimized on a population basis in other ways (*J*_*a*_, noise, *W* ^*e*^), or extracted from the average PET atlases (*R*_*D*1_, *R*_*D*2_, *R*_*Se*_) were fixed to values reported in Table S1.

**Figure S1:**
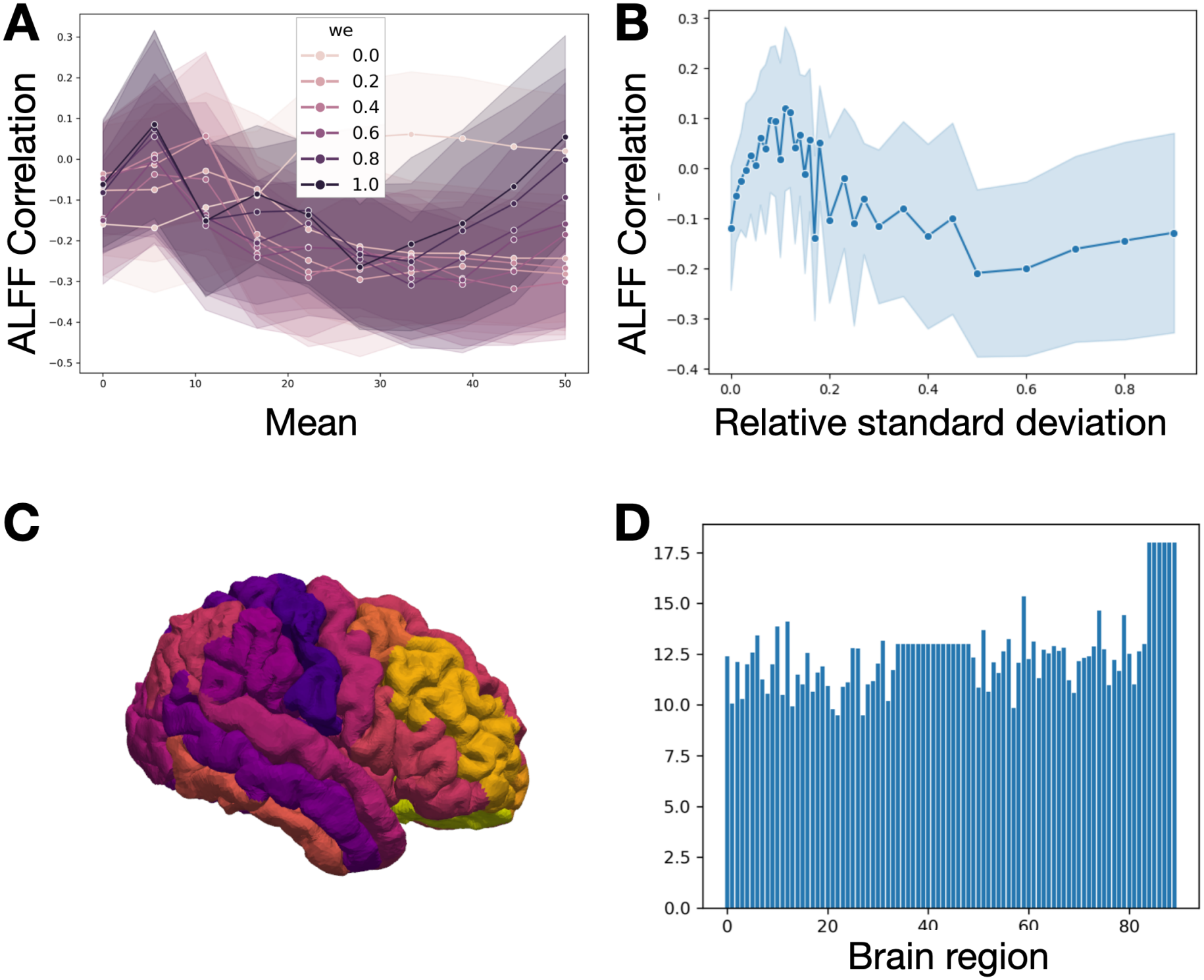
Parametrization of *J*_*a*_ and *J*_*sa*_. A: Relationship between the correlation of simulated and empirical cortical ALFF (y-axis) and the mean value of *J*_*a*_ (x-axis). Each line corresponds to a different value of global excitatory coupling (*w*^*e*^), while shaded regions represent variability across different standard deviations of *J*_*a*_. The correlation peaks around *J*_*a*_ = 11–12 for *w*^*e*^ in the range 0.2–0.4, which is the regime used in our simulations. B: Relationship between ALFF correlation (y-axis) and the relative standard deviation of *J*_*a*_ (defined as 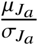). The correlation peaks at approximately 10%, indicating an optimal level of regional heterogeneity. Based on these results, we selected 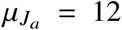 and 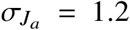 for subsequent simulations. C: Cortical surface visualization (Desikan–Killiany parcellation) showing regional z-scores of cortical gray matter volume, computed using CENTILE scores. D: Resulting *J*_*a*_ distribution used in simulations of the synthetic patient. Cortical values are derived from gray matter measures using Equations 15–16. Subcortical regions are assigned a fixed value (*J*_*a*_ = 12.5), while midbrain regions are set to a higher value (*J*_*a*_ = 17.5).

#### Receptor affinities of the medications

The baseline values of the Z parameters are adapted from (*63*), where the value corresponding to the D2 receptor is 2 to 4 times higher than those of the other two receptors, whose values are comparable. For each medication, we identify the receptor for which it exhibits the highest affinity. Starting from the baseline value of this receptor, we gradually reduce its Z parameter towards zero. Thus, the baseline value represents the absence of medication effects, whereas zero represents the maximum antagonist effect. The Z parameters of the other two receptors, for which the medication has lower affinity, are only partially reduced, proportionally to their affinity values normalized by the one of the receptor with the highest affinity. In this way, the largest effect is assigned to the receptor for which the medication has the highest affinity, while smaller effects are assigned to the other two receptors. The rescaling formula of *Z* for a drug at increasing doses is shown in Equation S1.

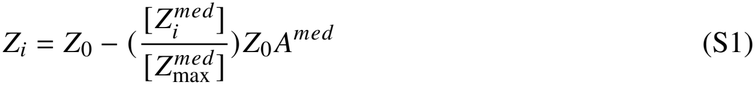

Here, *med* = {amisulpride, aripiprazole, clozapine, olanzapine}, *i* ∈ {0, …, 20}, and the normalized *A* value represents the normalized receptor affinity coefficient, with a value of 1 for the receptor with the highest affinity, as reported in Table 8. *Z*_*i*_ is the shifted Z parameter of a particular receptor, *Z*_0_ is its initial value under the effect of no medication (i.e., at 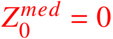). Because the shift of Z reflects increasing receptor occupancy (*O*_*i*_), a pharmacokinetic–pharmacodynamic framework described by (*27*) enables the conversion of the receptor occupancy (and therefore the simulated shift of Z) in clinically interpretable units of mg/day (*D*_*i*_), as shown by Equation S2 and S3.

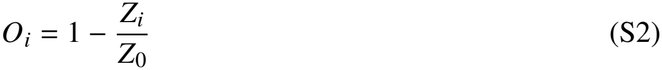

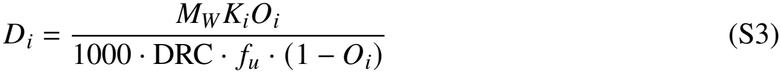

Here, *M*_*W*_ is the molar mass of the drug, Ki its receptor affinity, DCR the dose-to-receptor concentration conversion factor, *f*_*u*_ the unbound plasma fraction. For each drug, the conversion is based on the *Z* parameter associated with the receptor for which the drug exhibits the highest affinity. The resulting dose is capped at the maximum recommended daily dose. The corresponding values are reported in Table S1. We acknowledge that this approach represents a simplified approximation of pharmacological receptor modulation, as it relies on relative receptor affinity rather than explicitly modelling the distinct pharmacodynamic properties of each medication, such as their intracellular cascade activations. Nevertheless, it provides a reproducible and pharmacologically-grounded framework to link receptor occupancy, and therefore *Z*, to a clinically interpretable dose.

**Table S1:**
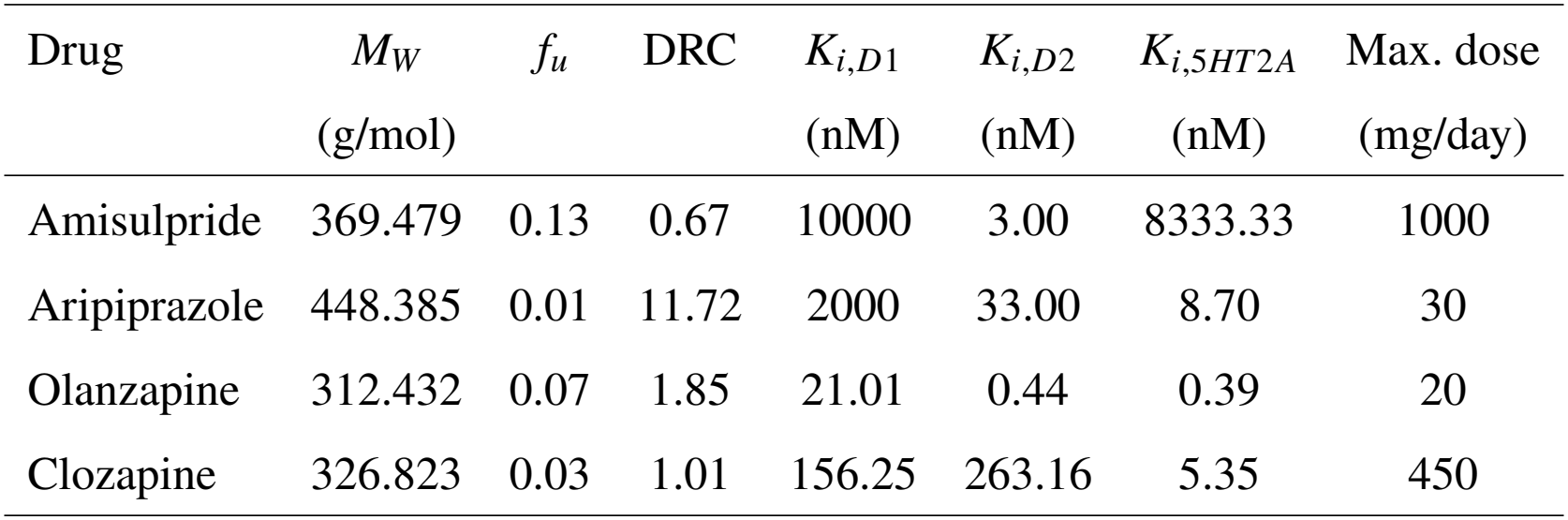
Pharmacokinetic and receptor affinity parameters used to estimate receptor occupancy as a function of administered dose.

| Drug | *M*<sub>*W*</sub> | *f*<sub>*u*</sub> | DRC | *K*<sub>*i*,*D*1</sub> | *K*<sub>*i*,*D*2</sub> | *K*<sub>*i*,5*HT*2*A*</sub> | Max. dose |
|---|---|---|---|---|---|---|---|
|  | (g/mol) |  |  | (nM) | (nM) | (nM) | (mg/day) |
| Amisulpride | 369.479 | 0.13 | 0.67 | 10000 | 3.00 | 8333.33 | 1000 |
| Aripiprazole | 448.385 | 0.01 | 11.72 | 2000 | 33.00 | 8.70 | 30 |
| Olanzapine | 312.432 | 0.07 | 1.85 | 21.01 | 0.44 | 0.39 | 20 |
| Clozapine | 326.823 | 0.03 | 1.01 | 156.25 | 263.16 | 5.35 | 450 |

#### List of extracted data features

In this work, we used 72 features extracted from either empirical or simulated BOLD signals. Two of these features—Fluidity and Global Brain Coupling (GBC)—are computed at the whole-brain level. Fluidity is defined as the variance of the dynamic functional connectivity (FCD) matrix, while GBC corresponds to the average value of the upper triangle of the whole-brain functional connectivity (FC) matrix. Regional functional connectivity (FC) is defined as the mean of the FC values in the row of the FC matrix corresponding to a given region. We considered 12 regions per hemisphere, resulting in a total of 24 regional FC features. The regional amplitude of low-frequency fluctuations (ALFF) is computed as the sum of the amplitudes of the Fourier-transformed BOLD signal within the frequency band of 0.01–0.08 Hz for each region. These ALFF values are then z-scored across all regions. Similarly, we included 12 regions per hemisphere, yielding 24 ALFF features. Finally, functional connectivity between specific pairs of regions is computed using the Pearson correlation between their BOLD time series. In total, we included 22 bilateral region pairs. For the computation of the mediation score, we selected Fluidity together with FC values from specific region pairs: Left Putamen–Caudal Anterior Cingulate Gyrus, Left Putamen–Rostral Anterior Cingulate Gyrus, Right Putamen–Caudal Anterior Cingulate Gyrus, Right Putamen–Rostral Anterior Cingulate Gyrus, Left Putamen–Insula, Right Putamen–Insula, Left Caudate–Hippocampus, and Right Caudate–Hippocampus. All features are listed in Table S3. Features used for the medication effectiveness score are highlighted in bold.

**Table S2:**
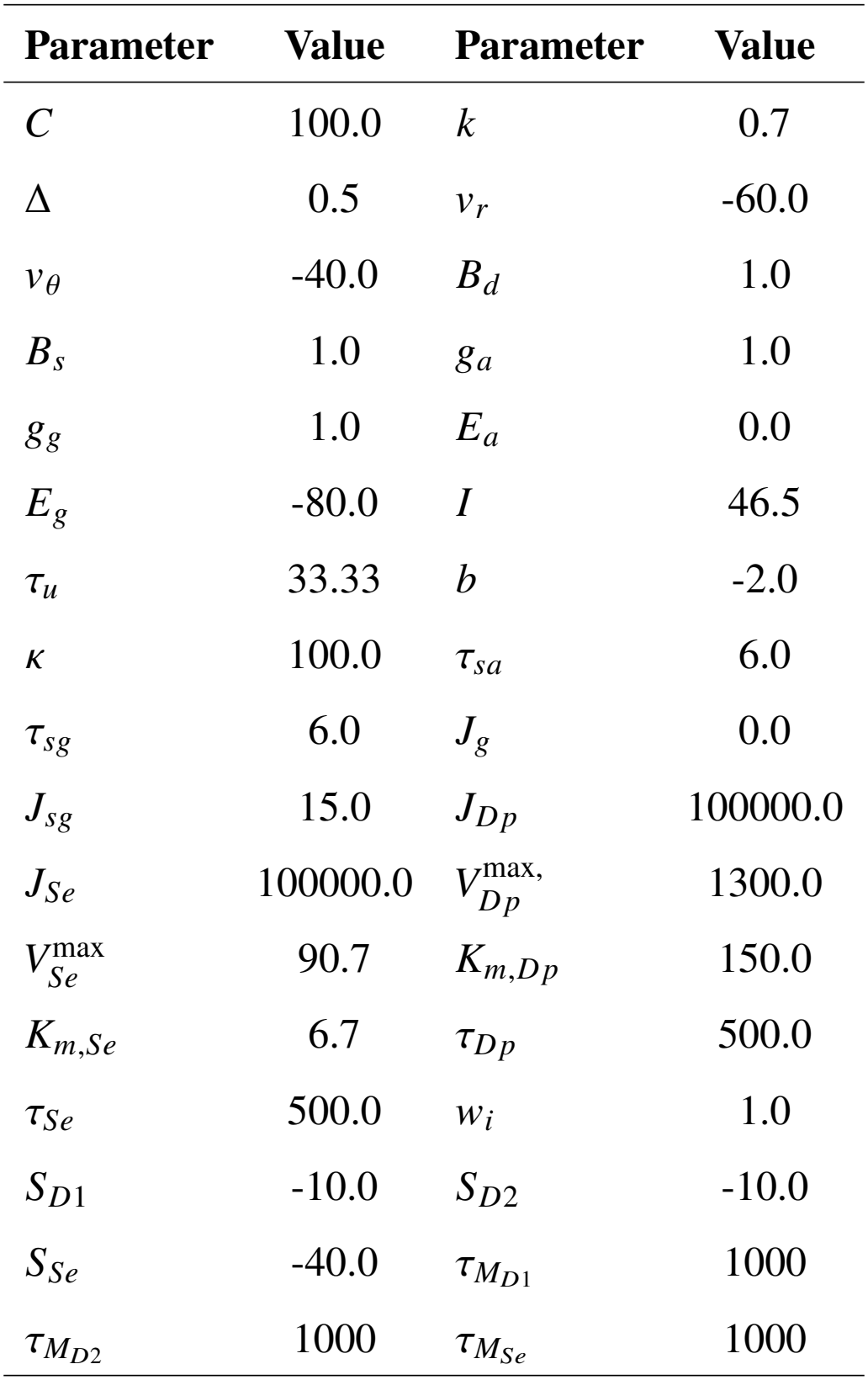
Default parameter values used in the model.

| **Parameter** | **Value** | **Parameter** | **Value** |
|---|---|---|---|
| *C* | 100.0 | *k* | 0.7 |
| Δ | 0.5 | *v*<sub>*r*</sub> | -60.0 |
| *v*<sub>*θ*</sub> | -40.0 | *B*<sub>*d*</sub> | 1.0 |
| *B*<sub>*s*</sub> | 1.0 | *g*<sub>*a*</sub> | 1.0 |
| *g*<sub>*g*</sub> | 1.0 | *E*<sub>*a*</sub> | 0.0 |
| *E*<sub>*g*</sub> | -80.0 | *I* | 46.5 |
| *τ*<sub>*u*</sub> | 33.33 | *b* | -2.0 |
| *κ* | 100.0 | *τ*<sub>*sa*</sub> | 6.0 |
| *τ*<sub>*sg*</sub> | 6.0 | *J*<sub>*g*</sub> | 0.0 |
| *J*<sub>*sg*</sub> | 15.0 | *J*<sub>*Dp*</sub> | 100000.0 |
| *J*<sub>*Se*</sub> | 100000.0 | *V*<sup>max,</sup><sub>*Dp*</sub> | 1300.0 |
| *V*<sup>max</sup><sub>*Se*</sub> | 90.7 | *K*<sub>*m*,*Dp*</sub> | 150.0 |
| *K*<sub>*m*,*Se*</sub> | 6.7 | *τ*<sub>*Dp*</sub> | 500.0 |
| *τ*<sub>*Se*</sub> | 500.0 | *w*<sub>*i*</sub> | 1.0 |
| *S*<sub>*D*1</sub> | -10.0 | *S*<sub>*D*2</sub> | -10.0 |
| *S*<sub>*Se*</sub> | -40.0 | *τ*<sub>*M*<sub>*D*1</sub></sub> | 1000 |
| *τ*<sub>*M*<sub>*D*2</sub></sub> | 1000 | *τ*<sub>*M*<sub>*Se*</sub></sub> | 1000 |

##### 4.7.1 Robustness analysis on the dopaminergic and serotonergic projections

Although scaled by their respective inferred *W*, dopaminergic and serotonergic weights and projections are imposed by PET densities and anatomical constraints, which can directly shape the SBI-inferred posteriors. To investigate the robustness of the model to variations in the connectivity structure, we performed an additional parameter sweep in which we introduced perturbations to the dopaminergic or serotonergic connections. We randomly sampled the perturbation of each connection from a normal distribution with an increasing standard deviation up to 50% of its original value. For each perturbation, we performed 20 simulations for 11 different values of the *W* scaling. This analysis allowed us to assess the consistency of the features extracted from the simulations under increasing changes in the imposed connectivity weights, and to verify that the model does not operate in a region of parameter space where even minor perturbations could lead to substantial changes in its dynamics. Results in S3 show the mean and standard deviation of three main data features averaged over the 20 simulations and 11 *W* values at increasing perturbation (”Error” on the x-axis). By increasing the perturbation, the average values of the extracted data features is stable, while their standard deviation increases linearly. If the model were operating in an unstable regime, sudden changes in the features would have appeared, which is not the case. In this way it is possible to verify that the model does not operate in a region of parameter space where even minor perturbations could lead to substantial changes in its dynamics, thus biasing the SBI results.

**Figure S2:**
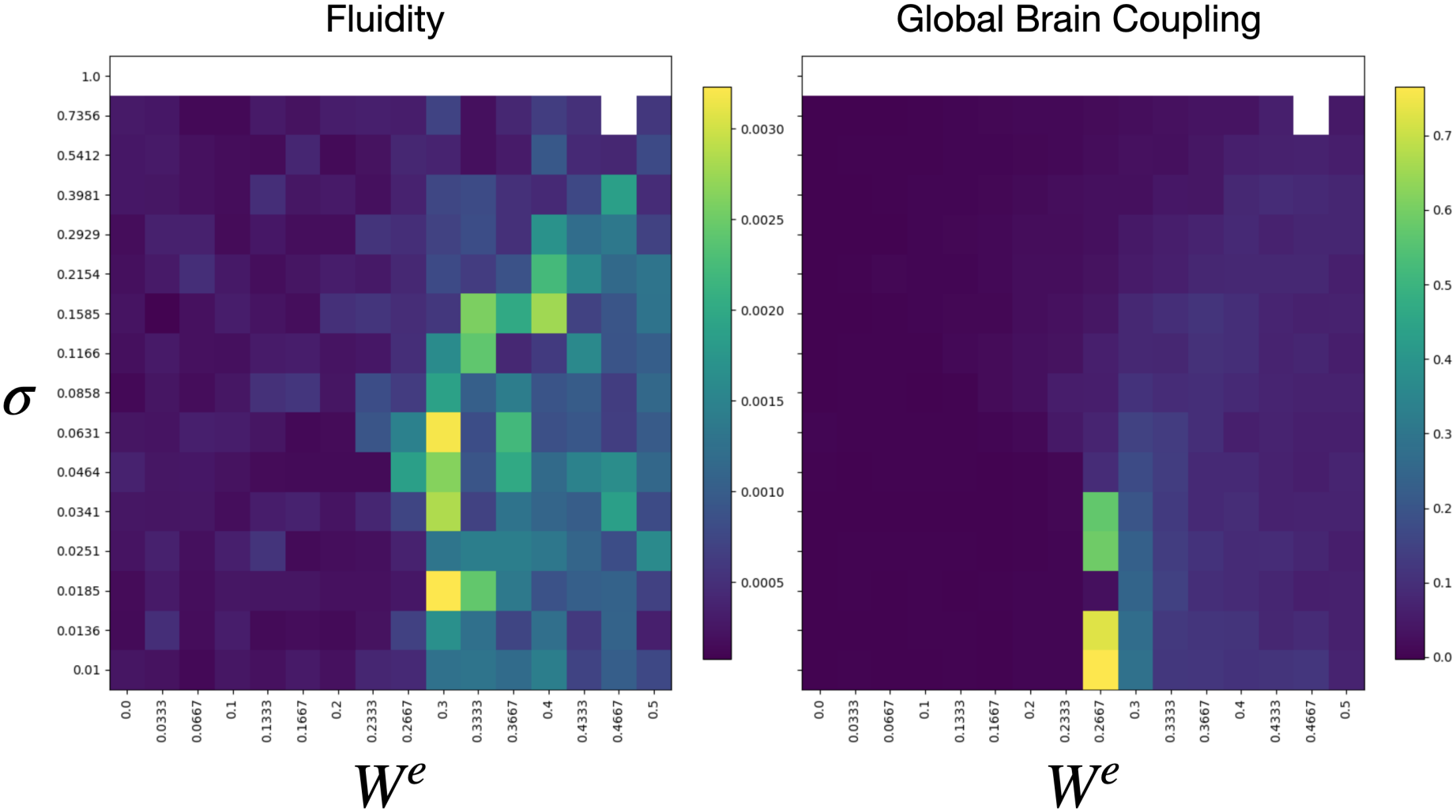
Parameter sweep over *W* ^*e*^ and *σ*. Heatmaps showing Fluidity and Global Brain Coupling in function of *W* ^*e*^ (on the x-axis) and *σ* (on the y-axis). It is possible to notice how both features have higher values at an intermediate range of *W* ^*e*^. Also note that for *σ* = 1.0, on top of the plots, simulations became unstable and it was not possible to compute the data features.

**Figure S3:**
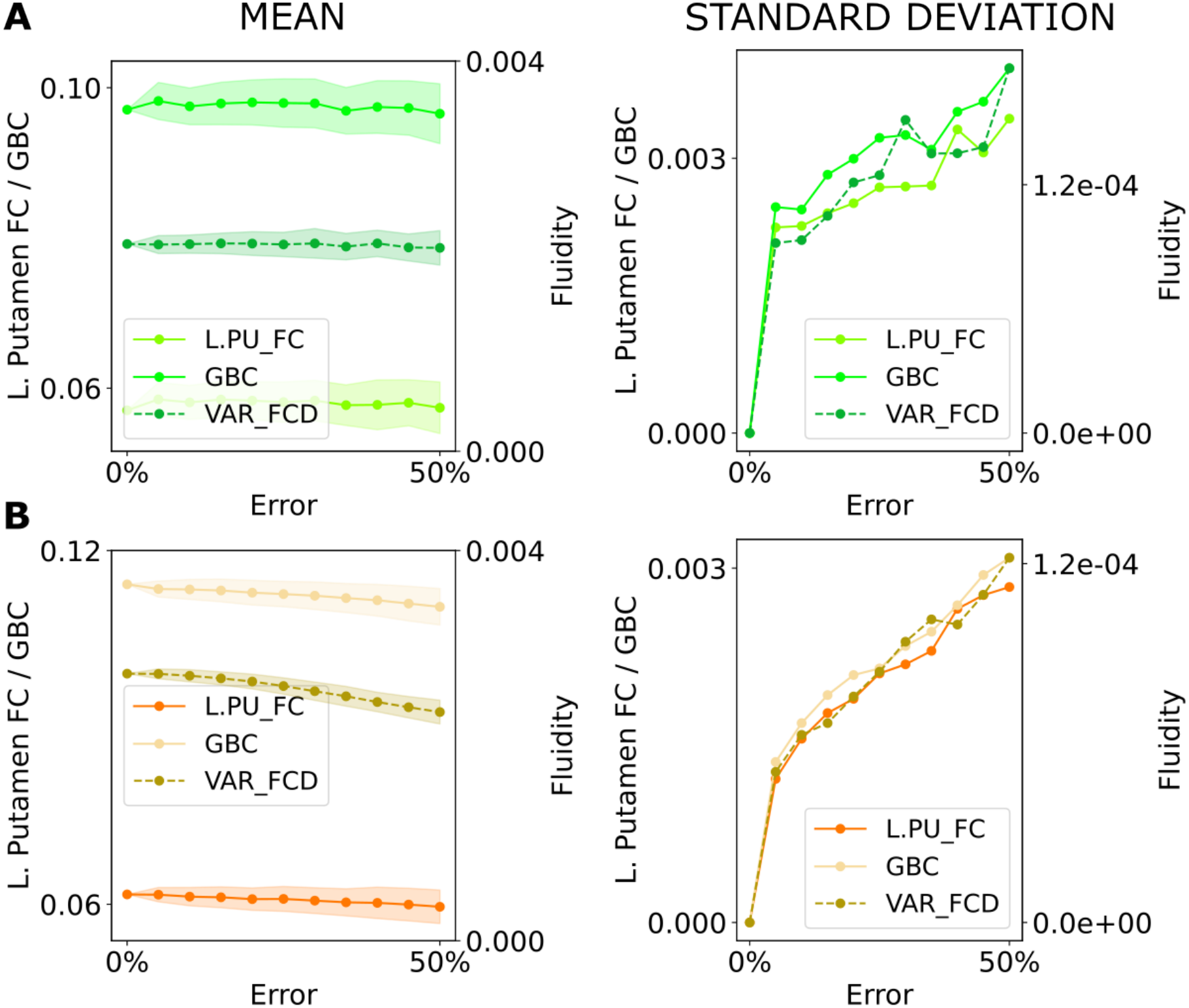
Robustness analysis on three data features. (**A**) Variation of the mean and standard deviation of the Left Putamen functional connectivity (L. Putamen FC), the mean of the functional connectivity (Global Brain Coupling, GBC), and Fluidity at increasing perturbation of the dopaminergic weights. The x-axis represents the standard deviation (“Error”) of the normal distribution from which the variability of each connection was drawn. It is possible to notice the stability of the mean of the three data features, while their standard deviation increases linearly. (**B**) Similar results for the perturbation of the serotonergic weights.

**Table S3:**
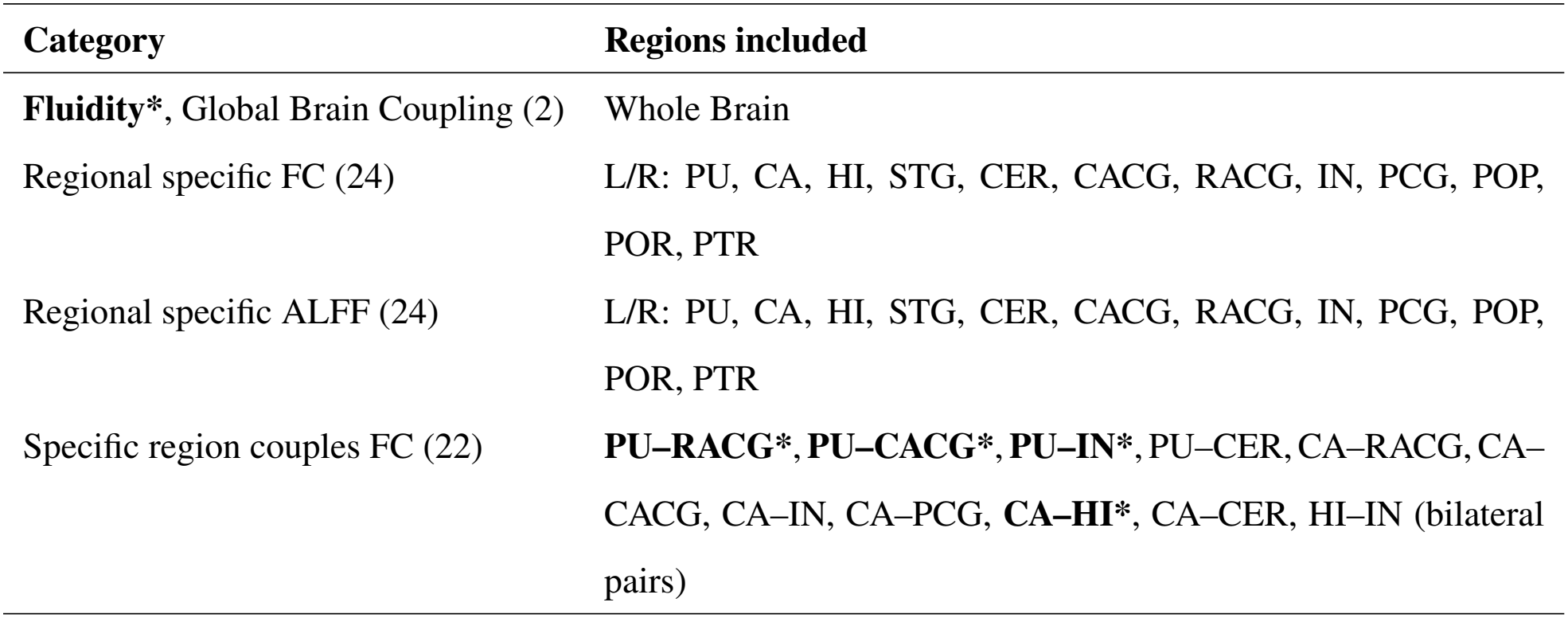
Summary of selected features grouped by category, regions, and computation method. FC = Functional Connectivity, ALFF = Amplitude of Low Frequency Fluctuations. Abbreviations (Desikan–Killiany atlas): PU = Putamen; CA = Caudate; HI = Hippocampus; STG = Superior Temporal Gyrus; CER = Cerebellum; CACG = Caudal Anterior Cingulate Gyrus; RACG = Rostral Anterior Cingulate Gyrus; IN = Insula; PCG = Posterior Cingulate Gyrus; POP = Pars Opercularis; POR = Pars Orbitalis; PTR = Pars Triangularis. L/R denote left/right hemispheres. Features for the medication score are highlighted in bold and by a *

| **Category** | **Regions included** |
|---|---|
| **Fluidity\***, Global Brain Coupling (2) | Whole Brain |
| Regional specific FC (24) | L/R: PU, CA, HI, STG, CER, CACG, RACG, IN, PCG, POP, POR, PTR |
| Regional specific ALFF (24) | L/R: PU, CA, HI, STG, CER, CACG, RACG, IN, PCG, POP, POR, PTR |
| Specific region couples FC (22) | **PU–RACG\***, **PU–CACG\***, **PU–IN\***, PU–CER, CA–RACG, CA–CACG, CA–IN, CA–PCG, **CA–HI\***, CA–CER, HI–IN (bilateral pairs) |

#### Medication threshold efficacy optimization on an initial cohort of 9 subjects

The threshold that the medication dose-response curve needs to cross to be considered effective by our pipeline was optimized to maximize the sensitivity, specificity and balanced accuracy of the classification of 9 initial subjects that were used as benchmark for the pipeline. We show in Fig. S4 the results of the sweep across increasing values of the threshold. We identified 0.1 as optimal threshold, where sensitivity is 80% (95% CI: 29%-99%), specificity is 75% (95% CI: 20%-99%) and balanced accuracy 77% (95% CI: 24%-99%). The binomial test yielded *p* ≈ 0.18.

**Figure S4:**
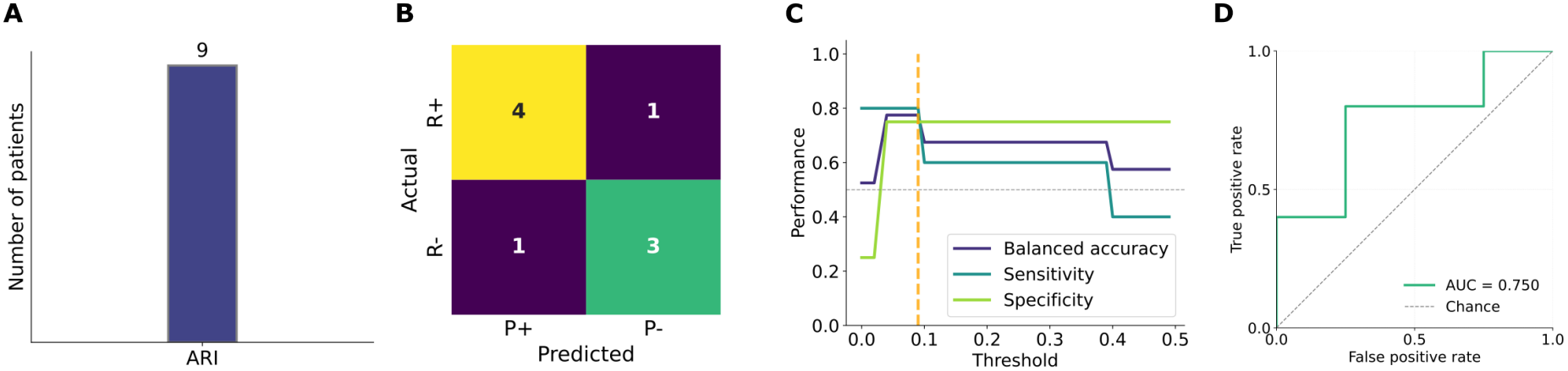
Accuracy of the pipeline prediction on a smaller cohort of 9 subjects. (**A**) Bar plot showing the number of patients receiving each antipsychotic as their last treatment at follow-up. (**B**) Contingency table showing the stratification of the 9 patients according to their empirical treatment response (actual responders, R+; actual non-responders, R−) and the pipeline prediction (predicted responders, P+; predicted non-responders, P−). (**C**) Sweep across threshold values showing the corresponding sensitivity, specificity, and balanced accuracy. The orange line indicates the optimized threshold. (**d**) ROC curve computed across increasing threshold values.

## Notes

### Competing Interest Statement

The authors have declared no competing interest.

### Author Declarations

Ethics committee/IRB of Aix-Marseille University waived ethical approval for this work.

### Summary of Updates

First revision round corrections. More patients were included in the analysis.

